# Does Contact Tracing Work? Quasi-Experimental Evidence from an Excel Error in England

**DOI:** 10.1101/2020.12.10.20247080

**Authors:** Thiemo Fetzer, Thomas Graeber

## Abstract

Contact tracing has been a central pillar of the public health response to the COVID-19 pandemic. Yet, contact tracing measures face substantive challenges in practice and well-identified evidence about their effectiveness remains scarce. This paper exploits quasi-random variation in COVID-19 contact tracing. Between September 25 and October 2, 2020, a total of 15,841 COVID-19 cases in England (around 15 to 20% of all cases) were not immediately referred to the contact tracing system due to a data processing error. Case information had been truncated from an Excel spreadsheet due to a row limit, which was discovered on October 3. There is substantial variation in the degree to which different parts of England areas were exposed – by chance – to delayed referrals of COVID-19 cases to to the contact tracing system. We show that more affected areas subsequently experienced a drastic rise in new COVID-19 infections and deaths alongside an increase in the positivity rate and the number of test performed, as well as a decline in the performance of the contact tracing system. Conservative estimates suggest that the failure of timely contact tracing due to the data glitch is associated with more than 125,000 additional infections and over 1,500 additional COVID-19-related deaths. Our findings provide strong quasi-experimental evidence for the effectiveness of contact tracing.

## 1 Introduction

The scientific and public debates around the most effective measures to mitigate the COVID-19 pandemic almost universally acknowledge the paramount importance of non-pharmaceutical interventions. Public health experts suggest that even after vaccines and treatments become available, such measures will remain necessary for a considerable amount of time (Ferguson et al., 2020). A central measure to contain COVID-19 has been the build-up testing-and-tracing capacities (Desvars-Larrive et al., 2020). The relative success of some countries in dealing with the pandemic has repeatedly been associated with the effectiveness of their COVID-19 tracing systems (Lu et al., 2020; Cowling and Lim, 2020; Anderson et al., 2020). Contact tracing comprises two key elements: first, people who have tested positive are contacted and asked to submit information on their recent, close contacts, and second, contact tracers attempt to reach each contact and encourage them to self-isolate for a period of time (WHO, 2020). This simple strategy has been a central pillar of communicable disease control in public health for decades. The eradication of smallpox in the 1970s, for example, is routinely credited to exhaustive contact tracing (Fenner et al., 1988).

Despite the simple appeal of contact tracing to contain the spread of infectious diseases such as COVID-19, significant doubts about its effectiveness remain (Clark et al., 2020; Li and Guo, 2020; Steinhauer and Goodnough, 2020). These revolve around two types of concerns. The first originates in the implementation of contact tracing: most systems rely on human contact tracers both to identify the contacts of infected persons and to get in touch with those contacts to impose or advise self-isolation. The success of contact tracing depends on the skills of contact tracers, who are often engaged at short notice and not well trained for their role (Manthorpe, 2020; Mueller and Bradley, 2020). Second, and more importantly, contact tracing may fail even if it is successfully implemented, because it fundamentally relies on eliciting the intended behavioral response. There is evidence that people may be unwilling to cooperate with contact tracers, and contact tracing faces opposition of various other kinds in the population. Infected person may not want to share complete or accurate information about their contacts, e.g., because they don’t take contact tracers seriously, distrust the government, are afraid of scams, social stigma, or have other privacy and cybersecurity concerns (Simko et al., 2020; Hakak et al., 2020; Cho et al., 2020; Altmann et al., 2020; Steinhauer and Goodnough, 2020). An even more pressing problem is that contacts may not believe contact tracers. This can lead to non-adherence to self-isolation, undermining the very purpose of contact tracing (Webster et al., 2020; Rubin et al., 2020). The obstacles faced by contact tracers have received significant media attention in the course of the COVID-19 pandemic.

Consider the case of England: an official report endorsed by the *Scientific Advisory Group for Emergencies* published on September 21, 2020, assessed the role of contact tracing, concluding that “[t]he relatively low levels of engagement with the system […] coupled with […] likely poor rates of adherence with self-isolation suggests that this system is having a marginal impact on transmission at the moment” (GOV.UK, 2020e). Notwithstanding this sobering assessment, the testing and tracing system in England was reported to draw on a budget of about *£*12 billion. Just in October 2020, over 1,100 additional Deloitte consultants were engaged to support the tracing system (Conway, 2020).

This striking disparity between, on one hand, the resources that countries around the world allocate to their contact tracing systems and, on the other hand, the uncertainty around their relevance to contain a pandemic call for a better understanding of their actual effectiveness. Policy evaluations are important because government interventions can have unintended consequences (Fetzer, 2020). The effectiveness of contact tracing, however, is notoriously hard to assess due to a lack of naturally occurring, exogenous variation. The existing literature therefore mostly relies on correlational evidence (Klinkenberg et al., 2006; Kendall et al., 2020; Kretzschmar et al., 2020b,a; Afzal et al., 2020; Kucharski et al., 2020; Park et al., 2020; Grantz et al., 2020). Correlational evidence on the role of contact tracing is subject to the concern that variation in the intensity of contact tracing across time or geographical areas is correlated with other variation not related to contact tracing, such as simultaneously occurring changes in other public health measures or area-fixed characteristics such as local populations’ different levels of adherence to self isolation.

In this paper, we exploit a unique quasi-experiment in England that generated exogenous variation in the intensity of contact tracing and allows us to estimate the causal effect of contact tracing on the evolution of COVID-19. England provides an ideal setting to study the effectiveness of contact tracing as it relies on a centrally managed testing-and-tracing system. On October 3, 2020, UK authorities announced that due to a “technical error,” 15,841 COVID-19 cases that should have been reported between September 25 and October 1 have not entered the official case statistics and were not referred to the central contact tracing system (GOV.UK, 2020d). Different areas in England were affected to very different degrees by the late referrals to contact tracing originating in the data glitch, providing our source of exogenous variation to determine the effectiveness of contact tracing. We will argue that the local affectedness by the technical glitch – caused by the 65,536 rows limit of the *XLS* file format used in the official data reporting process – is unrelated to any other factors that determined the previous spread of COVID-19. We conduct a series of analyses to test our identification assumption. These analyses strongly confirm that the local impact of the data glitch may indeed be random; specifically, it is unrelated to a large battery of area characteristics such the demographic makeup as well as to recent levels and trends in an area’s exposure to the COVID-19 pandemic as captured by a host of conventionally used outcomes.

15,841 missed referrals correspond to a share of 15-20% of all new cases reported in the time range under consideration. The resulting aggregate effects at the national level are illustrated in Panel A of Figure 1, which plots the daily number of positive cases that were officially published (red line) next to the total number of positive tests conducted at a given date (blue line). The evolution of these lines over time reveals the following patterns: (i) the red line trails behind the blue line prior to the data glitch, reflecting the “natural reporting lag” that occurs because tests need to be evaluated, processed and reported; (ii) the gap between the reported and actual cases widens drastically between September 20 and October 2, a mechanical result of the data glitch that also led to the spike in reported cases on October 3; (iii) we witness a strongly accelerated growth in the actual numbers of positive cases (blue line) during the time of the misreporting, which we will argue is the adverse effect of late referrals to contact tracing on pandemic growth; and (iv) we see persistently higher case numbers in the aftermath of the data correction, indicating that the data glitch propelled England to a different stage of pandemic spread.

**Figure 1:**
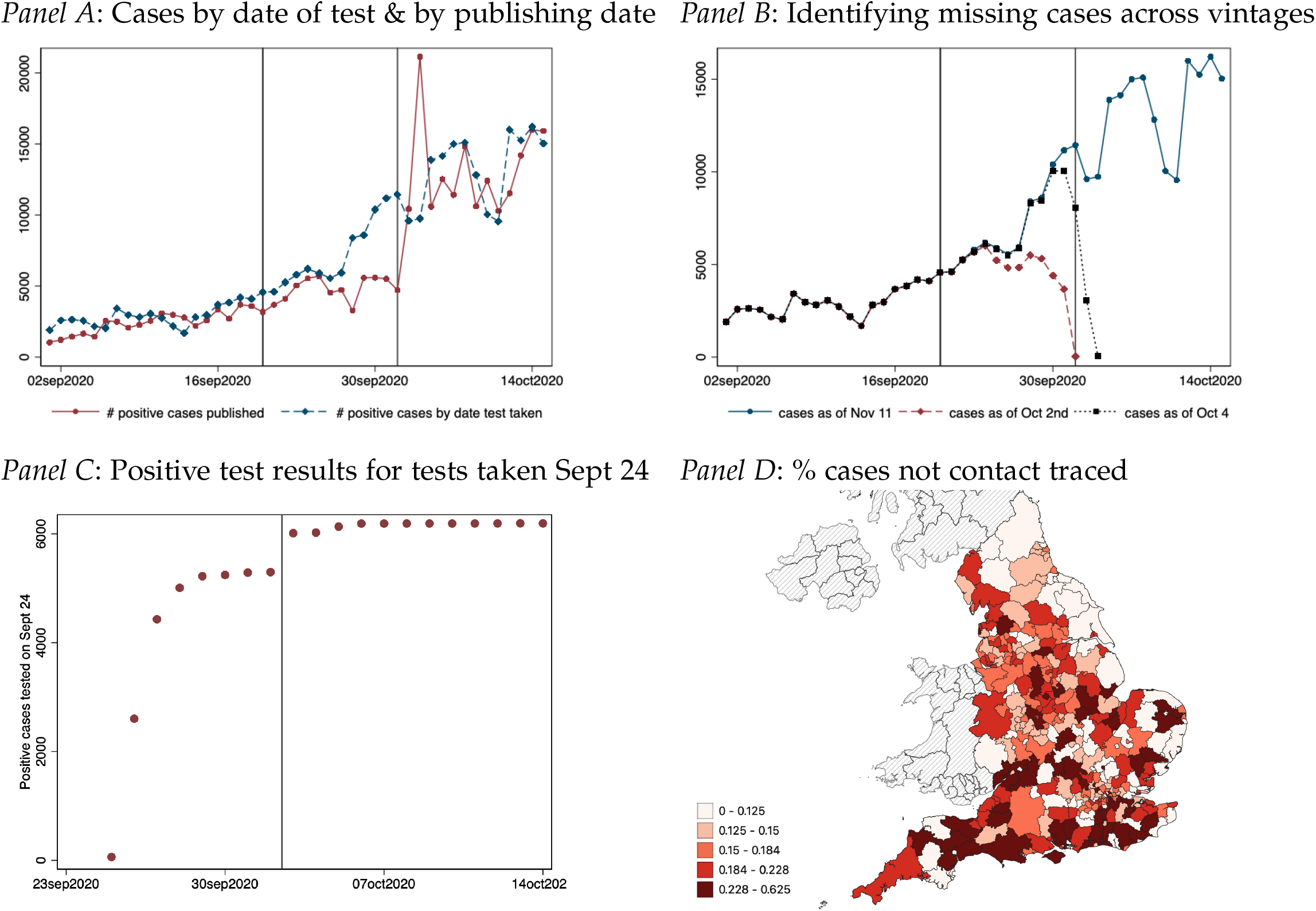
Delayed contact tracing referral: Identification of delayed referral to contact tracing. **Notes:** Panel A plots the number of positive cases based on the date that case results are published as well as the number of positive cases based on the date that the test was taken (not when the result was made public). There is a notable divergence between positive results published and positive test results from Sept 20 to Oct 2 capturing the delayed referral of positive cases to contact tracing. Panel B documents the number of cases by date on which a test was taken for three different versions of the dataset: Nov 11, Oct 4 and Oct 2nd. The data for Oct 4 includes a large set of the missing positive cases that were not reported in the Oct 2 data version resulting in large upward revisions. These revisions capture cases that were not referred to contact tracing until Oct 3 or 4th the earliest. Panel C illustrates this using data for all tests taken on Sept 24. Over time the reported value of positive COVID-19 cases converges to the true value as all test results get processed. Usually, 5 days after a test is taken at least 95% of all test results have been published. Between October 2 and October 3 the case count for Sept 24 jumps by around 715 cases or 12% of all cases due to the Excel glitch. Panel D illustrates the geographic distribution of the fraction of cases tested from Sept 20 to Sept 27 that were not referred to contact tracing until Oct 3 or Oct 4.

This paper leverages spatially and temporally granular data from England to investigate the potential causal effect underlying these observations. Our approach relies on a reconstruction, for each geographical unit, of the number of cases that were referred late to the contact tracing system as a result of the data glitch. This measure forms the basis for a series of difference-in-differences analyses to estimate the effect of contact tracing on variety of outcomes related to the spread of COVID-19. Our baseline estimations leverage a dataset containing 37,485 observations across 315 geographical units in England. We document robust and quantitatively large effects of contact tracing on the evolution of the pandemic. In areas with higher exposure to the contact tracing shock, we subsequently find a notable increase in COVID-19 infections and, with the usual delay, in COVID-19-related deaths. These effects go hand in hand with an increase in the test positivity rate, a sharp increase in number of tests performed and a worsening of the quality of contact tracing.

A large part of this paper is dedicated to examining the robustness of our findings. Among other things, we document that our results are robust (i) to different ways of constructing the measure of late referrals, (ii) to the level of spatial aggregation and (iii) the exclusion of individual regions, (iv) to different empirical strategies which compare areas that had been evolving very similarly in terms of a host of measures of pandemic development prior to the data glitch, but that were affected differentially by the data glitch itself, (v) to alternative functional forms of the estimated relationship (e.g., log-log specifications) that acknowledge the non-linear nature of infection dynamics, (vi) to controlling for a large vector of more than 50 additional area characteristics, (vii) to alternative ways of conducting statistical inferences, specifically, randomization inference, (viii) to controls for the effect that lower reported case numbers at the local level might have had on people’s behavior, (iix) and we conduct an empirically highly demanding placebo test.

Across this battery of analyses, we estimate that the specific failure of timely contact tracing due to the data glitch is associated with something between 126,836 (22.5% of all cases in the post-treatment period under consideration) and 185,188 (32.8%) additional cases, and with between 1,521 (30.6% of all deaths) and 2,049 (41.2%) additional COVID-19-related deaths. We advise caution, however, against taking these effect sizes at face value: due to the complex structure of a pandemic, such as significant externalities across areas and the non-linear nature of infectious developments, effect magnitudes are inherently difficult to interpret.

The rest of this paper is organized as follows. Section 2 provides background information on the context, data and the measurement approach. Section 3 describes the empirical strategy. Section 4 presents the results, discusses their significance and explores the robustness of our findings. Section 5 concludes.

## 2 Context and Data

### 2.1 Contact Tracing in England and the Data Glitch

In England, laboratories report positive COVID-19 test results to *Public Health England* (PHE) on a daily basis. The PHE aggregates all nation-wide test results using an automated reporting dashboard, which forms the basis for the official reporting of case numbers as well as contact tracing (GOV.UK, 2020a). Specifically, data on positive cases are passed on to the *NHS Test and Trace* (Test and Trace) system, a government-funded service that was established in 2020 to organize all contact tracing at the national level (GOV.UK, 2020c). For all cases that do not come from a high exposure setting such as a school or a prison, the infected person is contacted via a text, email alert or phone call and asked to shared details of their recent close contacts and places they have visited. They can respond online via a secure website or by telephone with a contact tracer. NHS initially employed a team of 25,000 contact tracers.

On October 4, 2020, the PHE released a public statement on a “technical issue” discovered in the night of October 2 to October 3 (GOV.UK, 2020d). An internal investigation had revealed that 15,841 positive cases had accidentally been missed in the data reported to Test and Trace. The maximum file size of 65,536 rows of the document type used for official data processing purposes (Excel Binary File Format, *XLS*) had been exceeded, leading to a failure to transfer case information in excess of that limit. PHE reported that the original reporting dates for these cases would have been between September 25 and October 2. While the data glitch did not affect people receiving their individual COVID-19 test results, an anticipated 48,000 close, recent contacts of COVID-19 patients had not been traced in a timely manner and had therefore not been encouraged to isolate (BBC, 2020). The evolution of the daily number of newly reported cases is depicted in Panel A of Figure 1. In the seven seven days preceding the discovery of the data glitch on October 3, newly reported cases averaged 4,853 per day, ranging from a low of 3,277 to a high of 5,599. The delayed reporting of some cases resulted in a severe jump in the daily case numbers: the officially reported number increased to 10,436 on October 3 and to 21,140 on October 4, before leveling off to an average of 11,814 reported new cases per day in the subsequent seven days. The Excel error attracted significant public and media attention.^1^

### 2.2 Data on COVID-19 in England

Our baseline analyses leverage three sources of publicly available data.

#### Reporting Dashboard

Our primary dataset is constructed using the UK’s COVID-19 dashboard.^2^ This dashboard provides granular data on COVID-19 infections and deaths at different spatial resolutions. Our geographical focus is on England, because other countries in the UK were not affected by the Excel error. The data include daily lab-confirmed positive test results and deaths. Data on positive cases are characterized by two dates: the specimen date, i.e., the date when the sample is taken from the person being tested, and the reporting date, i.e., the date when a positive case is first included in the published totals and referred to Test and Trace, so that contact tracing can begin. In order to reconstruct the time line of case reporting for each specimen date, we collect “vintage datasets” published on past reporting dates. The distinction between specimen and reporting date forms the basis for our analysis of late referrals due to the data glitch, as detailed in Section 2.3.

We conduct analyses at different levels of spatial disaggregation. England has 315 lower tier local authority districts (LTLA). While most COVID-19 data is published at this level, some data is only available at the upper tier authority district level (UTLA) – of which there are 149 in England. Our baseline analyses exploit variation at the LTLA level but we replicate our results at the UTLA level as well as NUTS3 region level, of which there are 93 units.

The resulting core dataset is a balanced daily panel. Our estimation window focuses on the period starting in calendar week 28 (starting July 6, 2020) all the way to calendar week 44 (starting October 26), covering a total of 37,485 observations.

#### Test and Trace Statistics

We also draw on data on testing and tracing statistics provided by NHS Test and Trace (GOV.UK, 2020b). These data are published weekly and provide some statistics on the effectiveness of the contact tracing efforts such as the fraction of contacts reached, delays as well as the total number of tests taken and test positivity rates. These data are available at different geographical and temporal resolutions than the daily case data. Specifically, while the COVID-19 test statistics are provided for the most granular lower tier local authority district level (LTLA), the contact-tracing data are more patchy and only available at the coarser UTLA level. The data are provided at the weekly level for weeks starting on Thursday and ending on Wednesday. This implies that calendar weeks are not cleanly separated in this dataset. We matched reporting windows to calendar week based on the largest overlap. For example, calendar week 39 ranges from September 21 to September 28. The nearest reporting window for the Test and Trace statistics is the week starting on September 24 and ending on September 30, which straddles four days of calendar week 39 and three days of calendar week 40. We match this week to calendar week 39. This implies, however, that the identification of the exact timing of effects is more challenging in the weekly data.

Lastly, we note that the data on contact tracing at the subnational level are far from complete. Data on the effectiveness of contact tracing is only available at the UTLA level. Various relevant pieces of information on the performance of contact tracing are not provided. For example, the data provided at the UTLA level are not providing information that is otherwise in more aggregated statistics, such as the time-taken for COVID-19 positive individuals to be reached; the time taken for those individuals to provide their close contacts information; the time taken for these contacts to be reached.

Appendix Figure A7 provides an overview of the process flow and highlights the sources of delays and potential caveats to bear in mind when studying and interpreting the data, especially relating to contact tracing and its performance. The performance of the system, for example, is undermined if e.g. a high fraction of COVID-19 positive individuals can not be contacted or reached. This naturally implies that also potential close contacts may not be identified. Similarly, even if an individual that tested positive is successfully contacted, they may not remember the individuals they spent notable time together during the time they may have been infectious. And, even if individuals provide details of close contacts, these may not be reached in a timely fashion or may not be reached at all.

There appears to be room for improvement in the comprehensiveness of public reporting and the statistical presentation of the data.^3^

#### Additional weekly death statistics

In addition to the daily death statistics, we also leverage weekly death statistics at the local authority level as published by the Office for National Statistics (ONS, 2020). These data report on new COVID-19-related deaths by the type of location where the death occurred, e.g., at home, in hospitals or in care homes. These data will be studied primarily as an auxiliary outcome data in the Appendix.

### 2.3 Identifying Delayed Test and Trace Referrals

We rely on granular data on positive COVID-19 tests to construct a measure capturing the extent to which positive COVID-19 cases have been affected by the delayed referral to contact tracing across different parts of England. The official PHE announcement only specified the total number of late referrals but provided no information about the geographical distribution and the specimen dates of these cases. A Freedom of Information request has been raised by the authors to obtain a detailed geographic and temporal break down of all cases that were referred to contact tracing with a delay – so far, these data have not been made available.^4^

#### Baseline measure of late referrals

Despite the lack of official data, we can infer which individual cases have been affected by a delayed referral. To do so, we study the reported case figures at different points in time. The logic of our approach follows from Table 1 and Panels B and C of Figure 1. Table 1 shows the COVID-19 case counts as they were reported on three different dates: November 15, October 4 and October 2. The case counts are broken down by the date on which the test sample was taken (specimen date). For all tests taken on September 24, the most recent figures from November 15 imply a total of 6199 positive cases known as of November 15. Because more than 1.5 months have passed between the specimen date of September 24 and the reporting date of November 15, all tests should have been processed and entered the statistics. We can interpret the number from November 15 as the final case count for this specimen date. In fact, the typical time lag between the specimen date and the reporting date is much shorter. Appendix Table A1 and Appendix Figure A3 highlight that usually, between 94% to 96% of all positive cases are identified and reported within the five days following the specimen date. For our baseline measure, we therefore restrict our attention to the earliest specimen dates that where likely impacted by the Excel error, September 20 to September 27. By the time the error was discovered on October 3, tests taken during this specimen date range should have almost fully entered the statistics under normal circumstances. Panel B of Figure 1 visualizes the striking discontinuity caused by the data glitch in the otherwise smooth increase of the fraction of cases reported in the days following a given specimen date. Taking the example of the specimen date September 24, we observe that the fraction reported had converged to a steady level by October 2, but then a sudden upward revision occurred on October 3. This stands in contrast to the overall smooth evolution of the fraction reported for specimen dates not affected by the data glitch, as shown in Appendix Figure A2. These figures capturing the over-time conversion to the final case count on a given specimen date leverage data from different historically published versions of the COVID-19 dataset.

**Table 1:** Measuring the Number of Missing Cases Across Data Set Vintages

| Date | PHE | *Cases by test date and vintage* |  |  | *$\Delta$ Missing case measure* |  |  |
| --- | --- | --- | --- | --- | --- | --- | --- |
|  |  | Nov 11 | Oct 2 | Oct 4 | Oct 4 - Oct 2 | Nov 11 - Oct 2 | Nov 11 - Oct 4 |
| 20 September 2020 | - | 4578 | 4308 | 4569 | 261 | 270 | 9 |
| 21 September 2020 | - | 4609 | 4540 | 4604 | 64 | 69 | 5 |
| 22 September 2020 | - | 5269 | 5102 | 5251 | 149 | 167 | 18 |
| 23 September 2020 | - | 5795 | 4962 | 5704 | 742 | 833 | 91 |
| 24 September 2020 | 957 | 6199 | 5297 | 6132 | 835 | 902 | 67 |
| 25 September 2020 | 744 | 5912 | 4732 | 5840 | 1108 | 1180 | 72 |
| 26 September 2020 | 757 | 5560 | 4048 | 5476 | 1428 | 1512 | 84 |
| 27 September 2020 | - | 5944 | 3635 | 5898 | 2263 | 2309 | 46 |
| 28 September 2020 | 1415 | 8405 | 4969 | 8317 | 3348 | 3436 | 88 |
| 29 September 2020 | 3049 | 8598 | 4793 | 8461 | 3668 | 3805 | 137 |
| 30 September 2020 | 4133 | 10400 | 3123 | 10040 | 6917 | 7277 | 360 |
| 01 October 2020 | 4786 | 11172 | 51 | 10073 | 10022 | 11121 | 1099 |

Notes: Table illustrates how the number of missing cases is identified contrasting different versions of case data published on the official English Coronavirus dashboard. We focus on our shock measure computing the missing cases between Sept 20 and Sept 26. The bulk of the increase in cases for tests taken between Sept 20 - Sept 27 is corrected by the data update between Oct 4 to Oct 2. Revisions of figures for Sept 20 - Sept 27 are marginal after Oct 4th.

Judging from the typical reporting lag as observed between September 1 and September 19, we would expect that at least 95.9% of the positive tests taken on September 27 and at least 99.3% of the positive tests taken on September 20 have been reported before October 3. In reality, however, this share turned out to be much lower as a result of the late referrals. Appendix Figure A3 visualizes the share of cases reported with different reporting delays – the fraction reported by day five following a specimen date dropped to roughly 60% during the period affected by the data glitch.

For reasons of parsimony, our baseline measure is constructed assuming that *all* cases taken between September 20 and 27 would have been reported by October 2 in the absence of the data glitch. Formally, this means we define the number of late referrals in district *i* that were likely due to the Excel error as

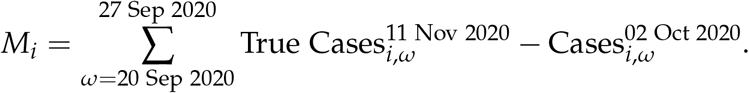

Specifically, across target specimen dates we sum up the difference between the final, “true” case count approximated by the most recent dataset version (November 15) and the case count known as of October 2. Note that, first, this baseline measure is transparent and does not impose auxiliary assumptions about the structure of the counterfactual reporting lag. Second, missed cases from earlier specimen dates are likely to have had the most pronounced effect on the development of the pandemic. A contact who contracted the disease from a person who tested positive on September 20, for example, could in turn infect others before the contact was finally traced by Test and Trace on October 3 or thereafter. This implies that the adverse effect of delayed contact tracing is stronger for cases with earlier specimen dates. In total, we calculate a number of 7,242 late referrals to contact tracing with specimen dates between September 20 and 27. This figure broken down to the LTLA level forms the basis for our measure of the local impact on late referrals due to the Excel error. We thus have a time-invariant scalar measure of treatment intensity in terms of late referrals.

Our baseline measure captures substantial variation in the extent to which different areas were affected by the data glitch. To illustrate, Panel D of Figure 1 shows a distinct geographic signature – there is substantial heterogeneity in the fraction of cases that we categorize as late referrals in each area. We confirm below that this heterogeneity is *as-if* random: it appears to be unrelated to all area-specific characteristics that are relevant for the local development of the pandemic, and we can therefore exploit it to evaluate the quasi-causal effects of the intensity of contact tracing.

#### Alternative measures of late referrals

While our baseline measure relies on just 7,242 late referrals out of the total of 15,841 cases that were officially acknowledged, our subset of late referrals from early specimen dates are likely to have had the strongest impact on the progression of COVID-19, and we can most cleanly identify these from the available sources of data. We construct a set of alternative measures of late referrals covering shorter or longer windows of specimen dates, e.g., from September 20 to 25, or from September 20 to 30. We further complement these analyses with a more parametric approach that statistically approximates the time path of the “typical reporting lag”, i.e., the distribution of delays in the absence of a processing error. Specifically, we proceed in two steps. First, for a given window of specimen dates, e.g., September 20 to October 1, we determine the reported case numbers that should be expected under the typical reporting lag, which we obtain by statistically approximating the usual evolution of reported fractions as shown in Appendix Figure A3. To this end, we estimate the fraction of cases that would be reported *d* days after the test was taken by fitting the following function using non-linear least squares:

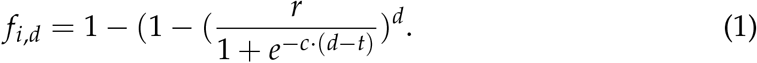

The above functional form is often invoked to approximate converging processes in variety of domains and can be estimated using non-linear least squares (see Steer et al., 2019 for an implementation in R). The fit of this estimation for the pre-treatment period of September 1 to September 19 is illustrated in Appendix Figure A5.

In a second step, we compare the predicted number of reported cases to the actually reported number of cases by October 2 to construct our measure of late referrals:

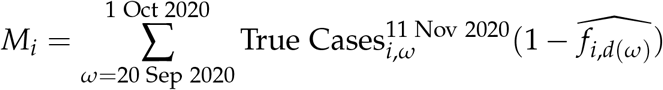

The above model 1 can be estimated at the country-level, but can also be trained at the region level to allow for region-specific variation in the typical reporting lag.

Our baseline specification relies on the number of late referrals normalized by the population size, while flexibly controlling for the local level and dynamics of the evolution of the pandemic. As an alternative measure, we can express the number of late referrals as a fraction of the final number of positive cases reported for September 20 to 27, by computing

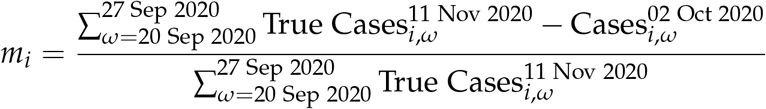

This measure is conceptually appealing in that it accounts for the local severity of the pandemic but it is statistically problematic due to a small sample issue. A fraction measure is noisy for areas with a low true case count, which creates a positive bias in our application. We use the above as an auxiliary measure imposing some sample restrictions, i.e., by focusing on places with at least a minimum number of cases. In the Appendix, we explore a variety of measures and show that our findings are robust to those.

## 3 Empirical Strategy

Our empirical strategy exploits cross-sectional variation in the extent to which different parts of England were affected by the delayed referral of COVID-19 positive cases to contact tracing efforts. This cross-area variation in exposure is quasirandom as a result of the Excel data entry error, allowing us to study the causal effect of contact tracing on measures of subsequent COVID-19 spread. We follow a simple and a refined difference-in-differences estimation approach at different geographic resolutions of the data.

### 3.1 Difference-in-differences specification

The basic difference-in-differences estimate is obtained from estimating

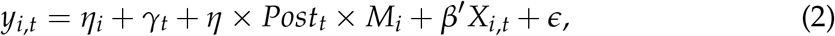

where *y*_*i,t*_ denotes a measure of COVID-19 spread in area *i* at time *t* (either a specific date or a week). The regression controls for district fixed effects, *η*_*i*_, as well as a set of time fixed effects *γ*_*t*_. To account for the non-linear nature of case growth we add a host of additional measures *X*_*i,t*_ of the disease progression across areas and control flexibly for these.

Specifically, we measure an area’s average number of new COVID-19 cases per capita, the number of tests per capita as well as the positivity rate of the tests during calendar weeks 37 to 38 (September 7 to September 20), directly preceding the data glitch. For each of these measures, we categorize districts into deciles according to its empirical distribution across districts. We successively control for non-linear time trends in these variables by decile. This ensures that we are not confounding or wrongly attributing differences in the outcome variables to the fact that different parts of England had been at different stages in the pandemic. Instead this specification aims at identifying the differential effect that late referrals to contact tracing had on the subsequent spread of COVID-19, comparing areas that have been on a very similar trajectory in the pandemic in the weeks just prior to the data glitch.

Naturally, the above exercise can be extended to flexibly estimate treatment effects over time. This will further allow us to shed light on the common-trends identification assumption implicit in the above difference-in-differences approach. Specifically, we estimate

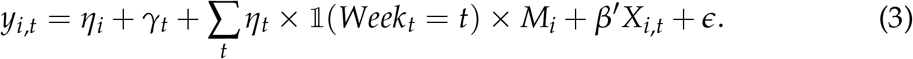

This allows us to plot the estimated coefficients 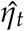 allowing us to explore to what extent differences in the outcome emerge around the time that the contact tracing shock happened and to what extent this affected the pandemic development going forward.

### 3.2 Zooming in on districts with similar pandemic evolution

We supplement our baseline difference-in-difference exercise with an additional exercise that aims to tackle potential concerns about the non-linear growth in cases. To do so, we refine the control group for our difference-in-differences design. For each district *i*, we compute the similarity between that district *i* and every other district *j* in terms of their disease progression just prior to the Excel error. To measure distance, we use the cosine similarity metric, applied to the following vector of seventeen characteristics **X**_**i**_ capturing the disease progression in a district *i*: new COVID-19 cases and deaths per capita reported on October 1, 2020; the number of COVID-19 tests along with the positivity rate in calendar week 40; the average number of new COVID-19 cases and deaths per capita in calendar weeks 37 and 38; the average number of COVID-19 tests per capita and positivity rate during weeks 37 and 38; and the growth in new COVID-19 cases, tests, deaths and the positivity rate between week 37 and 38. We also add measures of the pandemic progression in the first wave, such as the death rates in March to June, as well as other area characteristics, such as population density. The similarity measure is computed as:

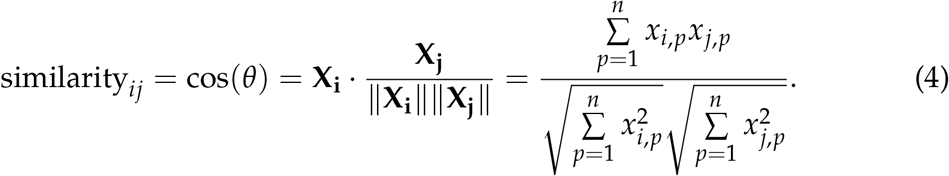

To illustrate this exercise, Appendix Figure A6 displays the cosine similarity measure between Adur and Watford (red diamonds), as well as their similarity to all other districts (blue dots) on the horizontal axis, along with a set of measures capturing the pandemic situation prior to the data glitch. Based on cosine similarity, Adur and Watford are closest to each other and are used to form a matched pair. Across all individual measures included in the similarity measure, the two districts are highly similar, showcasing that cosine similarity allows to identify districts with similar disease progression statistics. Even though the two districts share a cosine similarity score of 0.97, they differ substantially in terms of their number of late referrals, owing to the idiosyncratic effect of the data glitch. While Adur experienced 6.3 late referrals per 100k, Watford only saw 3.44 late referrals in that same time period. This highlights that our measure captures heterogeneity in exposure to the data glitch even in this matching approach that zooms in on otherwise highly similar districts.

For each district *i*, we identified a “best match” *j* using matching without replacement. We obtain 157 matched pairs from 314 districts, omitting the last district. We estimate a version of the above specification,

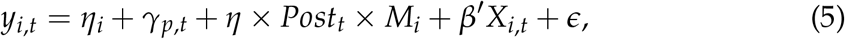

where we now control for matched-pair-by-time fixed effects *γ*_*p,t*_. In our most demanding specification, we control for 157 different sets of time fixed effects, allowing individual non-linear time trends for places that have been on a similar pandemic trajectory in the pre-treatment period. Practically, we estimate nearly 18,683 (157 × 119 days) separate time effects. This addresses the fact that infection dynamics in a pandemic may produce non-linear growth in cases. By virtue of zooming in on matched pairs of districts that look very similar in terms of the pandemic just around the Excel error occurred and affected these districts quite differentially, this will further strengthen the identification offered by this natural experiment.

## 4 Results and Discussion

We present our findings in four steps. First, we discuss the effects of late referrals on our main outcomes of interest, new infections and COVID-19-related deaths. Second, we begin to shed light on the corresponding mechanisms by looking at data on testing activity. Third, we zero in on the the impact of late referrals due to the data glitch on the performance of the Test and Trace system. In the fourth and final step of this analyses we present a battery of robustness analyses.

### 4.1 Effect on new infections and COVID-19-related deaths

Results from our main specifications are reported in Table 2. We begin by considering the effect on new infections. Results in column (1) of Panel A are obtained from estimating our main specification (equation (2)) using daily data at the LTLA level with 315 units (N=37,485). We regress the case count per capita on the number of late referrals per capita, controlling for time-and area-fixed effects as well non-linear time trends in measures of COVID-19 spread in calendar weeks 37 and 38, directly preceding the data glitch.

**Table 2:** Impact of non-timely contact tracing on the pandemic progression

|  | *plain difference-in-difference* |  |  | *matched pairs* |  |  |
| --- | --- | --- | --- | --- | --- | --- |
|  | (1) | (2) | (3) | (4) | (5) | (6) |
| *Panel A: Daily new COVID-19 cases per capita* |  |  |  |  |  |  |
| Post week 39 × Missing cases 20-27 Sept per capita | 0.606***<br>(0.111) | 0.619***<br>(0.111) | 0.606***<br>(0.119) | 0.514***<br>(0.120) | 0.560***<br>(0.119) | 0.586***<br>(0.121) |
| Mean DV | 11.790 | 11.848 | 11.906 | 11.810 | 11.869 | 11.927 |
| Observations | 28665 | 28301 | 28119 | 28574 | 28210 | 28028 |
| Spatial units | 315 | 311 | 309 | 314 | 310 | 308 |
| Additional controls | 1001 | 1911 | 2821 | 15288 | 16198 | 17108 |
| *Panel B: Daily new COVID-19 deaths per capita* |  |  |  |  |  |  |
| Post week 39 × Missing cases 20-27 Sept per capita | 0.008***<br>(0.003) | 0.008***<br>(0.003) | 0.007**<br>(0.003) | 0.006*<br>(0.003) | 0.006*<br>(0.003) | 0.005*<br>(0.003) |
| Mean DV | 0.183 | 0.184 | 0.185 | 0.183 | 0.184 | 0.185 |
| Observations | 28665 | 28301 | 28119 | 28574 | 28210 | 28028 |
| Spatial units | 315 | 311 | 309 | 314 | 310 | 308 |
| Additional controls | 1001 | 1911 | 2821 | 15288 | 16198 | 17108 |
| **Non-linear time trends in COVID-19 intensity weeks 36-38** |  |  |  |  |  |  |
| Cases per capita deciles x Date | X | X | X | X | X | X |
| Tests per capita deciles x Date |  | X | X |  | X | X |
| Positive test rate deciles x Date |  |  | X |  |  | X |

Our estimate for the difference-in-differences estimator that captures the daily increase in cases during calendar weeks 39 to 44 is 0.6. This implies that one additional late referral to Test and Trace was associated with a total of 25 additional cases over the subsequent six weeks. This implies that, given a total of 7,242 late referrals that we identified using our conservative baseline treatment measure, a total of 184,324 additional infections are likely associated with with the contact tracing delays due to the data glitch.

In all regressions, we non-parametrically control for non-linear time trends in infection dynamics as follows: we categorize districts into deciles according to a given measure of COVID-19 spread in the pre-treatment period (weeks 36 to 38) and include decile-by-date fixed effects accordingly. Next to non-linear infection dynamics based on total cases per capita (column (1)), we further allow for non-linear time trends across different pre-treatment testing intensities (column (2)) and the share of positive tests (column (3)). Our main estimate for the effect of late referrals on new cases (Panel A) is virtually unaffected by the different ways of accounting for infection dynamics.

Columns (4) to (6) present results for the refined difference-in-differences approach (see Section 3.2 in which we identify matched pairs of districts that went through a similar pandemic evolution prior to the Excel error. These results are discussed further in Section 4.4.

Figure 2 provides an an illustration of our regression results and sheds light on the role of area-specific trends in the pre-treatment period. Panel A displays the estimated weekly effects of late referrals due to the data glitch on the number of positive cases, obtained from estimating equation (3). Dates refer to specimen dates – i.e., the date on which a COVID-19 test was taken – of the corresponding cases. We make three observations. First, we do not find a significant relationship between our measure of late referrals and pre-treatment case numbers. Recall that the first reporting day affected by the data glitch was September 25, which due to the natural reporting lag impacted test results with specimen dates down to around five days earlier, i.e., September 19. This explains the small positive effect observed in calendar week 38 that ended on September 20. Second, we observe a pronounced positive effect of late referrals on case numbers in calendar weeks 39 to 41. The effect is largest in calendar week 40, roughly 2 weeks following the first COVID-19 cases whose contacts were not traced in a timely manner. The error was discovered and corrected late in week 40, on October 3. Third, the treatment effect of late referrals becomes statistically indistinguishable from zero from around calendar week 44, indicating that the impact of late referrals to contact tracing from late September had a notable, but temporary effect on infections over a four-week window.

**Figure 2:**
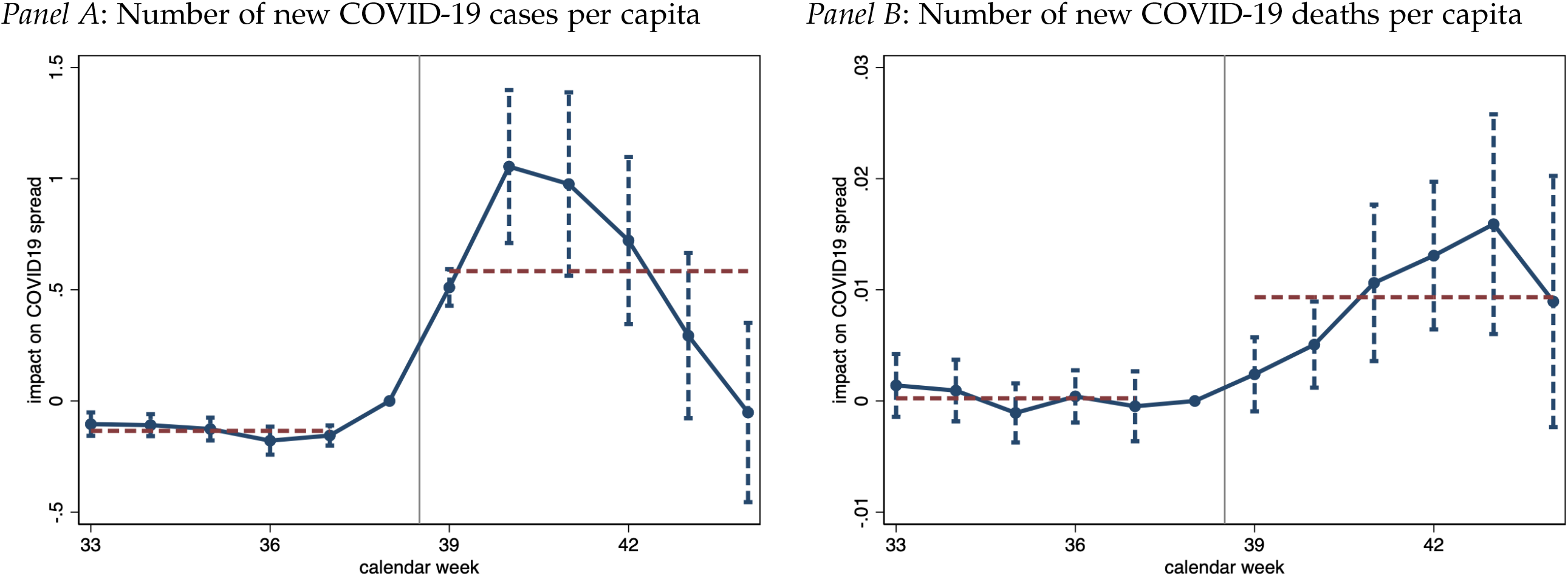
Impact of delayed referral to contact tracing, new COVID-19 infections and deaths. **Notes:** Figure presents regression estimates capturing the impact of cases that tested positive between Sept 20 to Sept 27 but were not referred to contact tracing until the earliest October 3, 2020 on the outcome variables indicated in the figure panel heads. All regressions control for district fixed effects and date fixed effects, along with non-linear time trends in the extent of true infections measured as of today during calendar weeks 37 and 38. Standard errors are clustered at the district level with 90% confidence intervals shown.

Analogous regression estimates for the effect on new daily COVID-19-related deaths are displayed in Panel B of Table 2. Our main difference-in-differences approach yields a precisely estimated and significant effect of 0.008 additional daily deaths per late referral to Test and Trace. This corresponds to 0.33 new COVID-19 deaths per late referral over the post-treatment time period studied (calendar weeks 39-44). From this, we estimate that the non-timely referral to Test and Trace resulted in an additional 2,433 COVID-19-related deaths in England. Comparing this number to the estimated increase in cases suggests that the implicit case fatality rate among additional COVID-19 infections due to the late referrals to contact tracing was around around 1.3%.

Panel B of Figure 2 illustrates the sizeable increase in deaths per capita that starts to become apparent from around calendar week 40. The effect becomes statistically significant at the 10% level by week 41, peaking in calendar week 43. The effect on the death toll lags behind the increase in cases shown in Panel A, which is due to the well-known lag between infections and subsequent deaths. The estimated effect is largest three weeks after the corresponding peak in the impact on positive cases and appears to subside by week 44, again following the trajectory of infections with a lag.

### 4.2 Effect on COVID-19 testing

We complement our baseline analyses by studying the mechanisms related to the effect of late referrals, leveraging different sets of outcomes. First, we investigate the impact on testing activity.

Contacts of infected persons are encouraged by contact tracers to self-isolate for 14 days. Note that without symptoms, a contact is neither required nor advised to take a test themselves (cf. NHS, 2020; GOV.UK, 2020c). A negative test does not rule out an infection and the procedures required to conduct tests can by themselves contribute to the spread of the disease if a person is already infectious. By contrast, a contact who is not reached by the contact tracing system and never learns of their potential infection will not self-isolate or take other precautionary measures, especially if they are asymptomatic or pre-symptomatic. A failure to trace contacts has two implications: First, affected contacts cannot respond to their potential infection, increasing the likelihood both of infecting others and of getting infected by a third infectious person if they are not already infected. Second, upon developing minor symptoms, contacts not reached by the tracing system are more likely to get a test in order to confirm their infection, which they might have been advised against by a contact tracer. As a consequence, holding everything else equal, more contact tracing may be associated with *less* testing overall.

In Panel A of Figure 3, we show the effect of late referrals due to the data glitch on the total number of tests taken in a given district, based on a regression specification analogous to the ones above (equation (2)). We document a sizeable increase in the number of tests conducted. The COVID-19 testing data is available at the weekly level. In order to be able to directly compare the magnitudes with the previous results, we divide the weekly testing figures by seven to obtain an estimate of the daily testing rate. Our main difference-in-differences estimate suggests that each late referral led to, on average, 2.7 additional tests taken per day between calendar week 39 and 44 (Table 3, Panel A, column (1)).

**Table 3:** Impact of non-timely contact tracing on weekly COVID-19 test data

|  | *plain difference-in-difference* |  |  | *matched pairs* |  |  |
| --- | --- | --- | --- | --- | --- | --- |
|  | (1) | (2) | (3) | (4) | (5) | (5) |
| *Panel A: Weekly Tests Per Capita* |  |  |  |  |  |  |
| Post week 39 × Missing cases 20-27 Sept per capita | 2.665***<br>(0.512) | 2.894***<br>(0.501) | 2.746***<br>(0.516) | 2.769***<br>(0.587) | 2.990***<br>(0.526) | 2.886***<br>(0.553) |
| Mean DV | 304.007 | 304.007 | 304.070 | 304.242 | 304.242 | 304.307 |
| Observations | 28301 | 28301 | 28119 | 28210 | 28210 | 28028 |
| Spatial units | 311 | 311 | 309 | 310 | 310 | 308 |
| Additional controls | 1001 | 1911 | 2821 | 15288 | 16198 | 17108 |
| *Panel B: Weekly Positive Tests Per Capita* |  |  |  |  |  |  |
| Post week 39 × Missing cases 20-27 Sept per capita | 0.674***<br>(0.122) | 0.685***<br>(0.122) | 0.676***<br>(0.130) | 0.560***<br>(0.135) | 0.617***<br>(0.135) | 0.648***<br>(0.138) |
| Mean DV | 13.468 | 13.468 | 13.496 | 13.493 | 13.493 | 13.521 |
| Observations | 27762 | 27762 | 27664 | 27671 | 27671 | 27573 |
| Spatial units | 311 | 311 | 309 | 310 | 310 | 308 |
| Additional controls | 1001 | 1911 | 2821 | 15274 | 16184 | 17094 |
| *Panel C: Share of tests returning a positive result* |  |  |  |  |  |  |
| Post week 39 × Missing cases 20-27 Sept per capita | 0.001***<br>(0.000) | 0.001***<br>(0.000) | 0.001***<br>(0.000) | 0.001***<br>(0.000) | 0.001***<br>(0.000) | 0.001***<br>(0.000) |
| Mean DV | 0.036 | 0.036 | 0.036 | 0.036 | 0.036 | 0.036 |
| Observations | 27762 | 27762 | 27664 | 27671 | 27671 | 27573 |
| Spatial units | 311 | 311 | 309 | 310 | 310 | 308 |
| Additional controls | 1001 | 1911 | 2821 | 15274 | 16184 | 17094 |
| **Non-linear time trends in COVID-19 intensity weeks 36-38** |  |  |  |  |  |  |
| Cases per capita deciles × Date | X | X | X | X | X | X |
| Tests per capita deciles × Date |  | X | X |  | X | X |
| Positive test rate deciles × Date |  |  | X |  |  | X |

**Figure 3:**
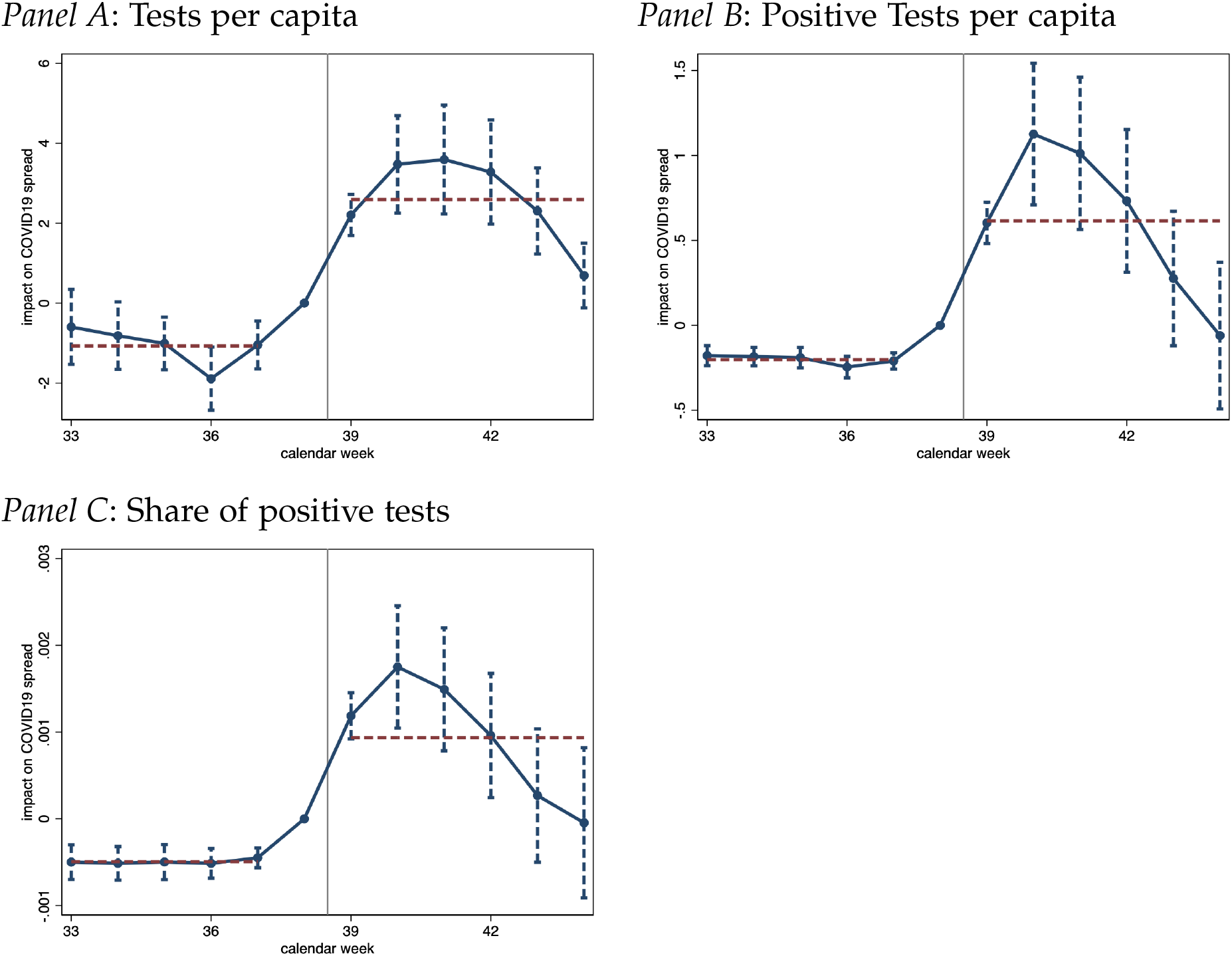
Impact of Delayed Referral of COVID-19 positive cases to Test & Trace on subsequent COVID-19 testing performance. **Notes:** Figure presents regression estimates capturing the impact of cases that tested positive between Sept 20 to Sept 27 but were not referred to contact tracing until the earliest October 3, 2020 on the outcome variables indicated in the figure panel heads. Note that the subnational Test & Trace statistics are made available lack a lot of detail and reporting is not following conventional calendar week definitions. Rather, a week refers to a time window ranging from Thursday to Wednesday of the subsequent week. That implies that the week 39 label, covering to the period from 24 Sep 2020 to 30 Sep 2020, straddles four days of calendar week 39 and three days of calendar week 40. All regressions control for district fixed effects and date fixed effects, along with non-linear time trends in the extent of true infections measured as of today during calendar weeks 37 and 38. Standard errors are clustered at the district level with 90% confidence intervals shown.

At the same time, we report a strongly positive effect on the number of positive tests per capita as well as the test positivity rate, see Panels B and C of Figure 3, respectively. We estimate that each additional late referral led to a significant increase in a district’s test positivity rate by 0.1 percentage points, given an average positivity rate of 3.6% (Table 3, Panel C, column (1)). The share of positive tests reverted back to pre-treatment levels in calendar week 43.

Our estimate of the effect on weekly positive tests per capita data lends credence to our above estimate for the effect on new infections which uses a different data source. We obtain a baseline estimate of 0.67 on positive tests (Table 3, Panel B, column (1)), which is closely in line and statistically indistinguishable to our estimate of 0.61 for new cases (Table 2, Panel A, column (1)).

These results underscore the intuition outlined above: because contacts not traced in a timely fashion did not change their behavior by self-isolating, and because they were more likely to take tests, we find that late referrals increased both the number of tests taken and the positivity rate.

### 4.3 Effects on the performance of contact tracing

Next, we analyze the repercussions on the effectiveness of the contact tracing system. To contain the further spread of the pandemic, a timely referral of cases to the contact tracing is essential. Unfortunately, the publicly available data on the Test and Trace system, especially on contact tracing performance, are far from exhaustive. As described in Section 2, contact tracing begins after positive cases are reported by laboratories to PHE, which in turn transfers case information to NHS Test and Trace (see also GOV.UK, 2020c). Contact tracers contracted by NHS Test and Trace then first contact individuals who tested positive. At this stage already, not all individuals that tested positive may be successfully reached. Even if an individual is reached and asked to provide contact details of recent close contacts, they may not properly recall or they may not be willing to disclose all relevant information. The actual contact tracing only sets in after contact information is obtained either through the contact tracer or the secure website. This implies that there are multiple margins through which contact tracing – even under normal circumstances – may fail: (a) not all COVID-19 positive individuals may be successfully reached; (b) those individual may imperfectly recall or incompletely disclose recent contacts; (c) and the contact tracing system may fail to reach all identified contacts.

Appendix Figure A1 studies aggregate performance data capturing the fraction of close contacts that were advised to self-isolate by the time taken to reach them. It demonstrates the possible effect that the data glitch had on the time taken to reach contacts. Note that this figure zooms in on the number of contacts that were actually reached, i.e., it focuses on step (c) above conditional on success in steps (a) and (b). While the fraction of those who were reached *within the first 24 hours* hovered above 80% in the weeks preceding the data glitch, the fraction plummeted to just above 60% in calendar week 40. Strikingly, we find that the tracing system’s performance remains low even in the three weeks following the correction of the data glitch. The share of contacts reached within 24 hours only appears to revert back to pre-treatment levels by week 44. This suggests that the tracing system was jammed by the late referrals from late September, adversely affecting the tracing performance for cases referred after the data glitch was corrected on October 3. Put differently, the tracing system may not have been well adapted to handle both a sudden influx of thousands of COVID-19 positive tested individuals referred to contact tracing with a delay and the subsequent higher infection levels that arose due to the preceding failure of a timely referral to Test and Trace.

In Figure 4 we present findings on the Test and Trace performance at the Upper Tier Local Authority level, analyzed using the same baseline specifications as above. Areas that experienced a larger impact on late referrals saw a deluge of referrals to contact tracing from calendar week 40. Note that a part of this increase may be mechanical as the contact tracing statistics for the week from October 1 to October 7 – that straddles calendar weeks 40 and 41, as explained in Section 2.2 – is matched to calendar week 40. We find that the impact on referrals persists throughout the subsequent weeks, similar to our estimates of the effects on infections and testing activity. This prolonged impact likely captures the fact that many of the individuals that were referred late to the contact tracing system further spread the disease, resulting in an overall worsening of the local pandemic situation.

**Figure 4:**
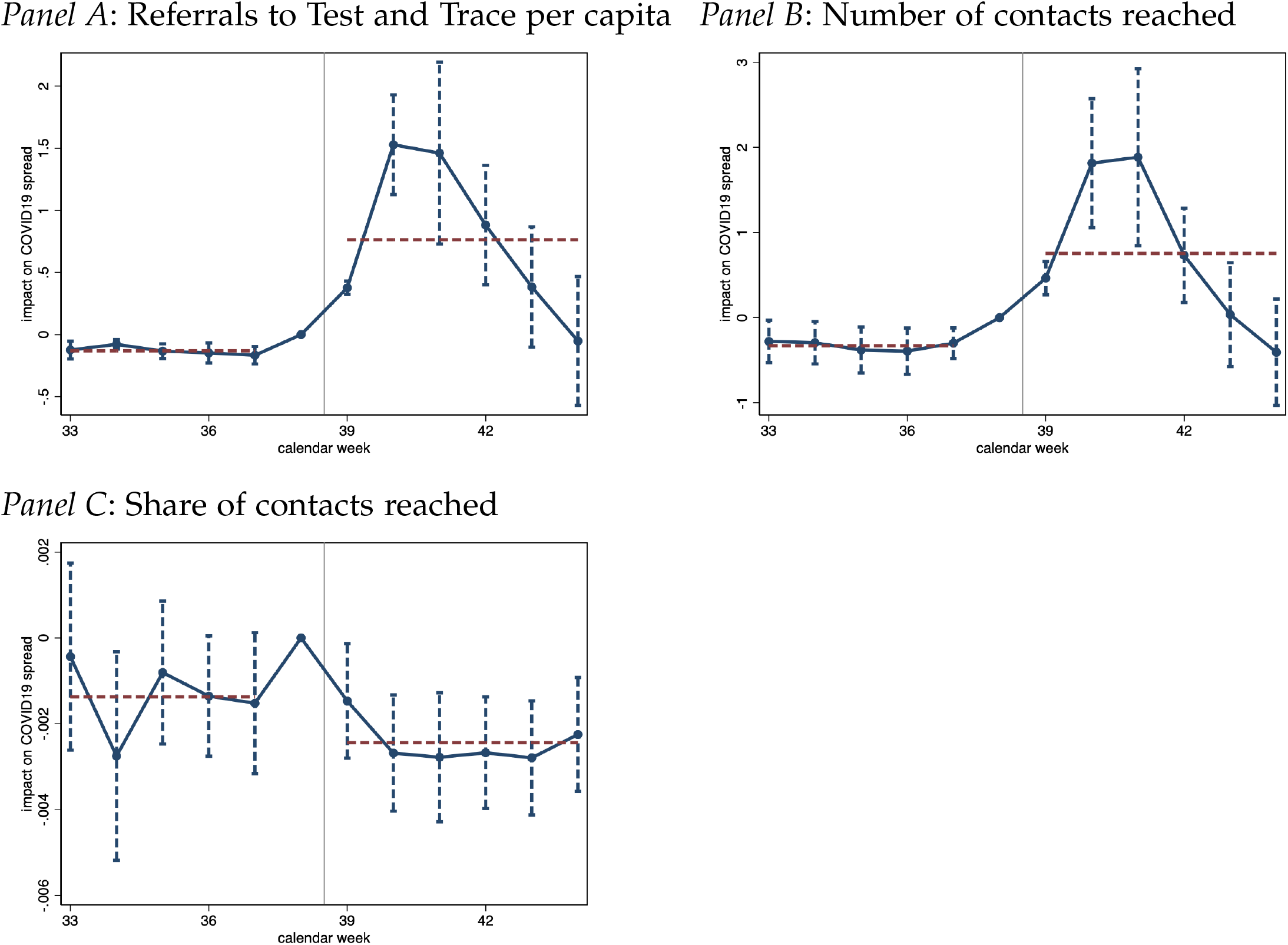
Impact of Delayed Referral to Contact Tracing on Test and Trace. **Notes:** Figure presents regression estimates capturing the impact of cases that tested positive between Sept 20 to Sept 27 but were not referred to contact tracing until the earliest October 3, 2020 on the outcome variables indicated in the figure panel heads. Note that the subnational Test & Trace statistics are made available lack a lot of detail and reporting is not following conventional calendar week definitions. Rather, a week refers to a time window ranging from Thursday to Wednesday of the subsequent week. That implies that the week 39 label, covering to the period from 24 Sep 2020 to 30 Sep 2020, straddles four days of calendar week 39 and three days of calendar week 40. All regressions control for district fixed effects and date fixed effects, along with non-linear time trends in the extent of true infections measured as of today during calendar weeks 37 and 38. Standard errors are clustered at the district level with 90% confidence intervals shown.

The only subnational performance measure available at the UTLA level captures the share of contacts reached out of all contacts recorded from those positively tested individuals who were both referred to the contact tracing system and successfully reached. These data do not include the time it took to reach individuals. In Panel C of Figure 4, we document some (more noisily estimated) evidence suggesting that the performance of contact tracing declined more drastically in parts of England that experienced a stronger impact on late referrals due to the data glitch. These estimates imply that the performance deteriorated with fewer close contacts being successfully reached. Late referrals are associated with a prolonged negative effect on the performance of the Test and Trace system that extends well beyond the correction on October 3. The corresponding difference-in-differences estimates are presented in Table 4.

**Table 4:** Impact of non-timely contact tracing on the weekly performance of contact tracing

|  | *plain difference-in-difference* |  |  | *matched pairs* |  |  |
| --- | --- | --- | --- | --- | --- | --- |
|  | (1) | (2) | (3) | (4) | (5) | (5) |
| *Panel A: Referrals to Test and Trace Per Capita* |  |  |  |  |  |  |
| Post week 39 × Missing cases 20-27 Sept per capita | 0.801***<br>(0.171) | 0.790***<br>(0.170) | 0.748***<br>(0.169) | 0.809***<br>(0.171) | 0.815***<br>(0.181) | 0.902***<br>(0.203) |
| Mean DV | 14.924 | 14.977 | 15.102 | 14.915 | 14.916 | 14.976 |
| Observations | 14378 | 14287 | 14105 | 14014 | 13832 | 13468 |
| Spatial units | 149 | 148 | 146 | 145 | 143 | 139 |
| Additional controls | 1001 | 1911 | 2821 | 7735 | 8554 | 9282 |
| *Panel B: Contacts Reached Per Capita* |  |  |  |  |  |  |
| Post week 39 × Missing cases 20-27 Sept per capita | 0.897***<br>(0.232) | 0.880***<br>(0.232) | 0.836***<br>(0.234) | 0.887***<br>(0.247) | 0.888***<br>(0.260) | 1.006***<br>(0.295) |
| Mean DV | 20.232 | 20.295 | 20.442 | 20.177 | 20.174 | 20.236 |
| Observations | 14378 | 14287 | 14105 | 14014 | 13832 | 13468 |
| Spatial units | 149 | 148 | 146 | 145 | 143 | 139 |
| Additional controls | 1001 | 1911 | 2821 | 7735 | 8554 | 9282 |
| *Panel C: Share of contacts reached* |  |  |  |  |  |  |
| Post week 39 × Missing cases 20-27 Sept per capita | -0.000<br>(0.001) | -0.000<br>(0.001) | -0.001<br>(0.001) | -0.002*<br>(0.001) | -0.001<br>(0.001) | -0.002*<br>(0.001) |
| Mean DV | 0.633 | 0.633 | 0.633 | 0.633 | 0.633 | 0.633 |
| Observations | 14378 | 14287 | 14105 | 14014 | 13832 | 13468 |
| Spatial units | 149 | 148 | 146 | 145 | 143 | 139 |
| Additional controls | 1001 | 1911 | 2821 | 7735 | 8554 | 9282 |
| **Non-linear time trends in COVID-19 intensity weeks 36-38** |  |  |  |  |  |  |
| Cases per capita deciles × Date | X | X | X | X | X | X |
| Tests per capita deciles × Date |  | X | X |  | X | X |
| Positive test rate deciles × Date |  |  | X |  |  | X |

### 4.4 Robustness exercises

We conduct a number of additional analyses to shed light on the robustness of our findings.

#### Refined difference-in-differences estimation using matched pairs

As outlined in Section 3.2, we construct a refined difference-in-differences estimator based on a procedure of matching areas which are highly similar in terms of their pre-treatment exposure to the pandemic. This approach aims at creating even more accurate treatment-control comparisons. In columns (4) to (6) of Table 2, we report results that correspond to those in columns (1) to (3) except for the different construction of control groups. We reliably estimate treatment effects that are statistically indistinguishable from the plain difference-in-differences approach. Similar robustness exercises are reported for the other outcome measures that we study, see columns (4) to (6) in Tables 3 and 4.

We point out that the matched-pairs design is empirically exceptionally demanding. By creating matched pairs and controlling for time-fixed effects specific to each pair, we conduct like-for-like comparisons by studying pandemic outcomes within pairs of districts that have been on a highly similar trajectory just prior to the data glitch.

#### Regional Heterogeneity

Before exploring to which extent our results are driven by individual regions, we examine the regional heterogeneity of the estimated treatment effects. To obtain these estimates, we refer back to our main difference-in-differences model and interact the main treatment measure with a set of region dummies, plotting out the coefficients along with 90% confidence bands. These are presented in Appendix Figure A8. The results suggest that the impact of delayed referrals on subsequent infections is most pronounced in the East Midlands, the North West, the South East as well as in Yorkshire. The effects on COVID-19-related deaths are more noisily estimated. This analysis suggests that the positive impact on deaths is most severe in the East, London, the North West and Yorkshire.

#### Sensitivity to geographic regions and spatial aggregation

So far, we reported our analyses at the the level of the Lower Tier Local Authority (315 units, Table 2). In Appendix Tables A2 and A3 we replicate our findings at the Upper Tier Local Authority Level (149 units) as well as the NUTS3 region level (93 units). Moreover, in Appendix Figure A9 we examine the sensitivity of our findings to excluding individual areas from the estimation. We show the distribution of the leave-one-out-estimator of the effect of late referrals on new cases and deaths, separately for analyses conducted at the LTLA, UTLA and NUTS3 levels. The observed sensitivity of the treatment effects to excluding individuals regions is small.

#### Alternative functional forms for the relationship between late referrals and COVID-19 spread

Our main regression specifications estimate the effect of the per capita level of late referrals on the per capita level of measures of COVID-19 spread, controlling for the level as well as non-linear trends in pre-treatment exposure to the pandemic. The non-linear nature of infection dynamics suggests specifications with logarithms as an alternative. In Appendix Table A4, we additionally estimate the same type of regressions using different combinations, such as a log-log as well as a log-levels specification, replicating our main findings.

#### Alternative measures of late referrals

As outlined in Section 2.3, we made conservative assumptions to construct our baseline measure of late referrals, but there is some degree of flexibility in the calculation of the treatment measure. We explore the sensitivity of our findings to the use of alternative approaches in Appendix Table A6. Our main measure of late referrals aggregates all cases with a specimen date between September 20 and September 26 that were not referred to Test and Trace as of October 2. This measure is conservative in terms of the number of late referrals it predicts: it relies on 7,242 late referrals that can most clearly be identified as such, which is less than half of the officially reported figure of 15,841 late referrals. We report regression results analogous to those in Table 2 for three alternative ways of inferring of late referrals that are due to the data glitch.

To this end, we non-parametrically estimate the time path of the *typical* reporting lag from the time immediately preceding the data glitch, i.e., for specimen dates between September 1 and September 20 as described in Section 2.3. This allows us to predict the fraction of cases with a given specimen date that should be reported a given number of days after the test was taken.

We use this prediction exercise, first, to create an even more conservative measure than our baseline by subtracting the number of cases that we would expect to not have have been reported by October 2 under the typical reporting lag. As argued above, this barely affects our measure. Even for the latest date in the specimen date range considered, September 27, we would expect 95.9% of cases to have been reported by October 2 under normal circumstances (see Table A1). This more conservative measure reduces our predicted number of late referrals from 7,242 to 6,044.

Second, we create a more comprehensive, yet less conservative measure by including specimen dates of up to October 1, and by accounting for the typical reporting lag using the same non-parametric estimation as above. Note that due to the potential divergence between the estimated typical reporting lag from pre-treatment data and the actual reporting lag, this measure is noisy, especially for lower spatial aggregation. This measure leads to a total number of 9,755 late referrals, still below the officially reported figure of 15,841 missed cases.

Third, we re-run our analyses using our baseline measure of late referrals as a fraction of the total number of cases between September 20 and September 27 in a given area. As discussed in Section 2.3, this measure suffers from statistical bias: because a fraction measure is noisy in areas with low case counts, it has an artificial upward bias. We present estimation results for the fraction measure as well as a version that exclude areas with a total case count below 50 during the time of September 20 and 27.

All of these results are presented in Appendix Table A6. We replicate our main results for each of these three alternative measures, and more compellingly, find that the estimated effect magnitude varies little across specifications.

#### Placebo tests

In Appendix Figure A10, we present a series of placebo tests. To do so, we construct a simple estimator of late referrals by specimen date date as follows. First, for each specimen date, we retrieve the number of cases that was reported as of seven days following that specimen date. Second, we subtract this number from the final, “true” case count for that specimen date, which is the number of cases known for this specimen date as of the most recent version of the data. This allows us to construct, for each specimen date, a measure of the number of cases that are were not yet reported as of one week following the specimen date. We construct this measure for each specimen before, during and after the period that was affected by the Excel error.

The hypothesis of this placebo exercise is that judging from this measure of late referrals for a specific specimen date, only specimen dates between September 20 and September 25 should be predictive of future case growth. For tests taken on September 26, the case count seven days was already subject to the correction of the data glitch that occurred on October 3.

We test this hypothesis by running our main specification (equation (2)) using these measures. The results are presented for both new infections and new COVID-19-related deaths in Appendix Figure A10. We document that only the missing case figures constructed in this fashion from around September 20 to September 26 strongly predict subsequent case growth and deaths.

#### Additional controls

We conduct an exercise to address concerns about potential confounding factors or non-linear trends in *other* area characteristics that may be amenable to affect the spread of COVID-19. In Appendix Table A5, we re-run our main analyses on new infections and deaths while inculding additional controls. In columns (2) and (4), we add a large vector of 55 additional area characteristics and interact them fully with a set of time fixed effects. The area characteristics are: employment shares in 1-digit industries; educational attainment; socio-economic status of the resident population, which also captures shares in full time education or in university; and regular in-, and out commuting flows. These characteristics come from the 2011 Census. We also leverage the detailed demographic makeup of an area’s population by expressing population demographics as shares in ten year age intervals. We further control for death rates in the first wave of the pandemic in spring 2020; population density and its variability across small geographies within an area. Throughout, despite these empirically highly demanding specifications that control for non-linear case growth that may be induced by, e.g., school- or university reopenings, the results remain virtually unchanged.

#### Alternative inference

Inference in the paper is conducted using clustering of standard errors at the spatial level at which the outcome data is measured. An alternative is to conduct a type of randomization inference. To do so, we draw repeated random samples of the main missing cases measure, redistributing the missing cases randomly across districts. We do this in three ways: reshuffling district exposure measures *M*_*i*_ across all districts in England; across all districts within the 9 NUTS1 regions; and across all districts within the 33 NUTS2 regions. For each exercise, we create 100 reshuffled treatment exposure measures using these three approaches. This allows us to estimate the treatment effects for these placebo treatment assignments. We would expect that the point estimate that are obtained based on the true spatial distribution of the missed cases to be sharply different from the null effects we would expect for the reshuffled distribution.

The latter may not be the case, especially for the reshuffling exercises at the region or NUTS2 region level: due to potential spatial autocorrelation, our treatment effect estimates may spuriously pick up treatment effects due to such spatial correlation. We present these results for new COVID-19 cases and deaths in Appendix Figure A11 as a set of kernel density plots of the distribution of the 100 point estimates that are obtained from these placebo exercises. We indicate with a vertical line the point estimate obtained from using the true distribution of the district exposure measures *M*_*i*_. Throughout the exercises and the outcomes, we can reject the null hypothesis that the effect we observe is spurious with implied *p*-values that are below 0.01%.

#### The behavioral effect of lower reported case numbers

Our analyses so far emphasize the effect that the delays in contact tracing have played for the progression of the pandemic. Note, however, that the data glitch simultaneously led to lower publicly announced new case numbers. Local variation in the share of cases that was missing from these announcements might have affected people’s behavior. More specifically, lower local case counts may be associated with less social distancing, more public activities etc. It is unclear to which extent the population attends to and internalizes case numbers at the the local level. The most salient figures are arguably those at the national level. The aggregate national growth in cases due to the data glitch, however, is orthogonal to the regional variation that our analyses exploit. To assess to which extent this endogenous response to reported local cases numbers affects our results, we employ a method of approximating people’s activity based on mobility data (Google, 2020), as has been successfully done in other work related to COVID-19 (Fetzer, 2020; Fetzer et al., 2020; Yilmazkuday, 2020). Previous research showed that mobility measures pick up people’s response to COVID-19 such as staying at home and predict the progression of the pandemic. As shown in columns (3) and (6) of Appendix Table A5, however, controlling for a mobility metric does not affect estimates of the effect of late referrals, indicating that the behavioral response to the local number of reported cases does not play a significant role here.

### 4.5 Quantification of effects

We offer a tentative quantification of the effects across the whole of England and the English regions in Table 5. We anchor these point estimates on the main point estimates presented in Table 2 as well as the most conservative point estimate obtained from our most saturated specification in Appendix Table A5.

**Table 5:** Quantification of impact of delayed or missing contact tracing on pandemic spread between calendar weeks 39 to 44

| *Panel A: Cases* |  | Implied estimates |  |  |  |  |  | Ranges |  |  |  |
| --- | --- | --- | --- | --- | --- | --- | --- | --- | --- | --- | --- |
| Region | Cases | Col (1) Table 2 |  |  | Col (3) Appendix Table A5 |  |  | Levels |  | % of all cases |  |
|  |  | Estimate | 90% CI |  | Estimate | 90% CI |  | Low | High | Low | High |
| North East | 44878 | 16573 | 11559 | 21587 | 11404 | 6828 | 15979 | 6828 | 21587 | 15% | 48% |
| North West | 161232 | 66903 | 46661 | 87145 | 46036 | 27565 | 64506 | 27565 | 87145 | 17% | 54% |
| Yorkshire & ... | 101873 | 27164 | 18945 | 35382 | 18691 | 11192 | 26190 | 11192 | 35382 | 11% | 35% |
| East Midlands | 59919 | 12703 | 8860 | 16547 | 8741 | 5234 | 12248 | 5234 | 16547 | 9% | 28% |
| West Midlands | 63546 | 17922 | 12500 | 23345 | 12332 | 7384 | 17280 | 7384 | 23345 | 12% | 37% |
| East of England | 29047 | 6339 | 4421 | 8257 | 4362 | 2612 | 6112 | 2612 | 8257 | 9% | 28% |
| London | 61371 | 19017 | 13263 | 24771 | 13085 | 7835 | 18336 | 7835 | 24771 | 13% | 40% |
| South East | 43307 | 10845 | 7564 | 14126 | 7462 | 4468 | 10456 | 4468 | 14126 | 10% | 33% |
| South West | 32208 | 7128 | 4972 | 9285 | 4905 | 2937 | 6873 | 2937 | 9285 | 9% | 29% |
| England | 597381 | 184595 | 128745 | 240445 | 127018 | 76056 | 177981 | 76056 | 240445 | 13% | 40% |

| *Panel B: Deaths* |  | Implied estimates |  |  |  |  |  | Ranges |  |  |  |
| --- | --- | --- | --- | --- | --- | --- | --- | --- | --- | --- | --- |
| Region | Deaths | Col (4) Table 2 |  |  | Col (6) Appendix Table A5 |  |  | Levels |  | % of all deaths |  |
|  |  | Estimate | 90% CI |  | Estimate | 90% CI |  | Low | High | Low | High |
| North East | 694 | 209 | 86 | 331 | 136 | 26 | 246 | 26 | 331 | 4% | 48% |
| North West | 2195 | 842 | 348 | 1337 | 550 | 107 | 992 | 107 | 1337 | 5% | 61% |
| Yorkshire & ... | 1085 | 342 | 141 | 543 | 223 | 43 | 403 | 43 | 543 | 4% | 50% |
| East Midlands | 653 | 160 | 66 | 254 | 104 | 20 | 188 | 20 | 254 | 3% | 39% |
| West Midlands | 747 | 226 | 93 | 358 | 147 | 29 | 266 | 29 | 358 | 4% | 48% |
| East of England | 474 | 80 | 33 | 127 | 52 | 10 | 94 | 10 | 127 | 2% | 27% |
| London | 529 | 239 | 99 | 380 | 156 | 30 | 282 | 30 | 380 | 6% | 72% |
| South East | 547 | 137 | 56 | 217 | 89 | 17 | 161 | 17 | 217 | 3% | 40% |
| South West | 272 | 90 | 37 | 142 | 59 | 11 | 106 | 11 | 142 | 4% | 52% |
| England | 7196 | 2324 | 960 | 3689 | 1516 | 295 | 2738 | 295 | 3689 | 4% | 51% |

Notes: Table provides a quantification exercise of the implied effects of the delayed contact tracing of COVID19 positive cases on subsequent infections and deaths across English regions from calendar week 39 to 44 inclusive. Cases and Deaths refers to the cumulative total of new COVID19 infections and deaths since week 39 up to week 44 inclusive. The subsequent columns provides the estimate of the number of cases and deaths that appear econometrically linked to the cases that have not been referred to contact tracing. The table provides on the figures implied by the central point estimate as well as the most conservative estimate. It further provides ranges associated with 90% confidence intervals for the individual point estimates. The column head makes a reference to the specific point estimates leveraged.

To arrive at the presented figures, we leverage the point estimate and simulate the full distribution of effects for the post-treatment period that ranges from calendar week 39 to including calendar week 44. For the cumulative new infections, our point estimates suggest that with 90% confidence, between 13% to 40% of the nearly 600,000 new *detected* COVID-19 infections may be attributable to the failure to contact tracing. This calibration implies that 127,018 infections, or around 21% of all detected infections may be due to the contact tracing failure.

The numbers of additional COVID-19-related deaths linked to the error are estimated less precisely. Our central conservative point estimate would suggest that, out of the total of 7,196 COVID-19 deaths during the time window, a similar share of around 21% are due to the contact tracing error.

The table provides a range of further upper- and lower-bound estimates as implied by the 90% confidence intervals spanning around the point estimates. It also highlights that, not surprisingly, the effect is quite homogenous across the English regions in a relative terms.

We advise caution, however, against taking these effect sizes at face value: due to the complex structure of a pandemic, such as externalities across areas and the non-linear nature of infectious developments, effect magnitudes are inherently difficult to interpret.

## 5 Conclusion

The large existing literature evaluating the effectiveness of contact tracing exploits correlational relationships (see, e.g., Klinkenberg et al., 2006; Kendall et al., 2020; Kretzschmar et al., 2020b,a; Afzal et al., 2020; Kucharski et al., 2020; Park et al., 2020; Grantz et al., 2020). This paper paper contributes to this line of work by providing a causal estimate that leverages a unique source of quasi-experimental variation. The gist of our main findings squares with the previous state of evidence: despite numerous challenges faced by a contact tracing system such as a population’s potential lack of trust, non-adherence and privacy concerns, this non-pharmaceutical intervention can have a strong impact on the progression of an infectious disease. In the context under consideration, the non-timely referral to contact tracing due to a data blunder has likely propelled England to a different stage of COVID-19 spread at the onset of a second pandemic wave. Our most conservative estimates imply that the data glitch is directly associated more than 125,000 additional infections and over 1,500 additional COVID-19-related deaths.

We acknowledge the following limitations of this study. First, there is substantial heterogeneity in how different countries organize and implement their contact tracing systems. Similarly, there is likely to be high variation in the extent to which the population at difference places responds to and cooperates with the official test-and-trace efforts. The results obtained in our analyses should thus be viewed in the specific context of England with a nationally centralized tracing system. We do not suggest any generality of our findings for other countries. Second, the currently available data does not permit an exact reconstruction of the specimen dates of cases that were referred late to Test and Trace. As a consequence, we made highly conservative assumptions in our identification of late referrals. The number of late referrals that can be identified with high confidence as used in our baseline analysis (7,242) strongly understates the officially reported figure of late referrals (15,841). This implies that, in all likelihood, our conservative approach underestimates the total impact of the data glitch.

## Data Availability

All data and code used to produce the results presented in the paper will be made available upon publication of the manuscript.

### Appendix

**Figure A1:**
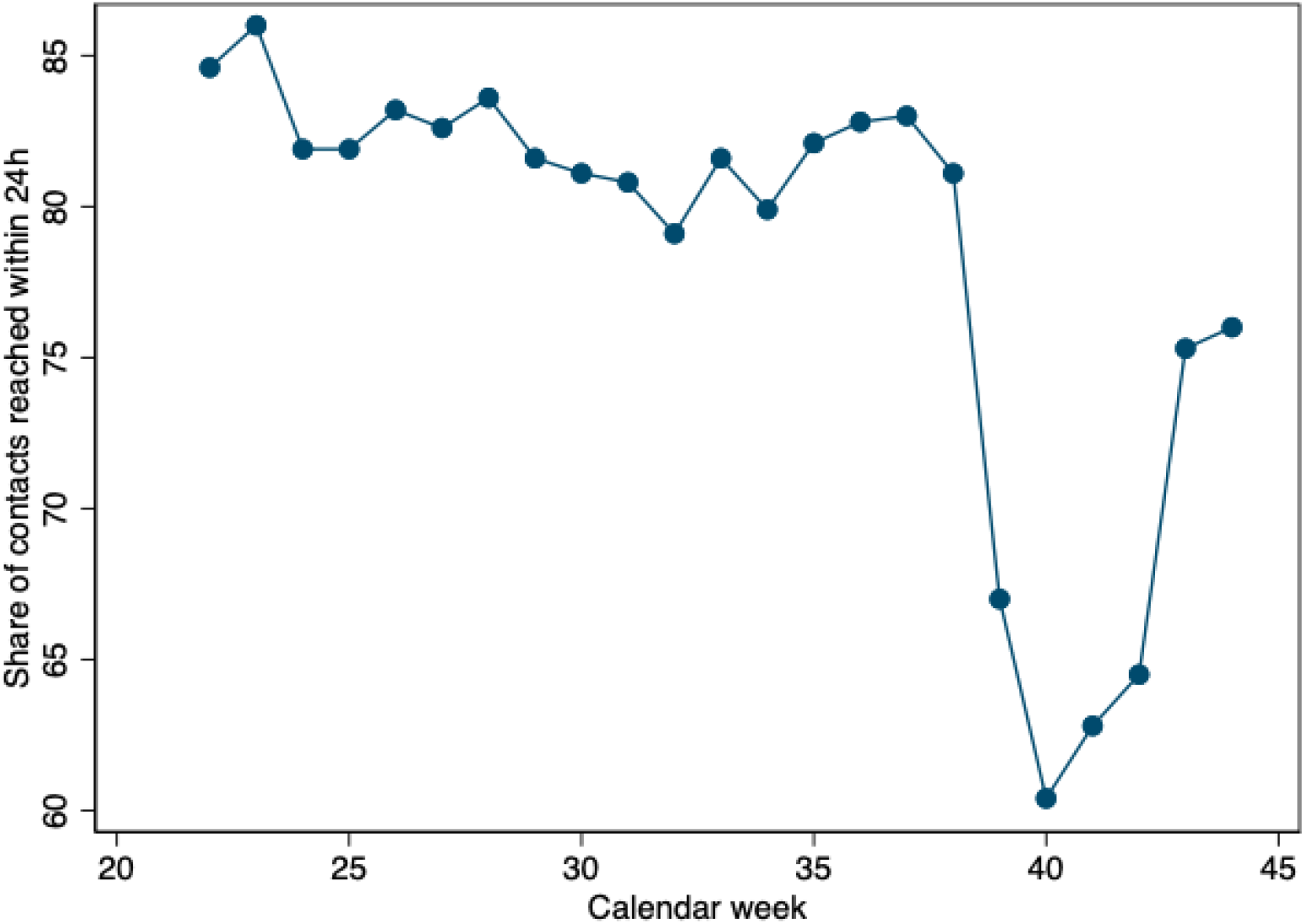
Evolution of performance of centrally managed contact tracing effort over time. **Notes:** Figure plots the share of contacts of individuals who were advised to self-isolate by time taken to reach them. The vertical axis presents the share of all contacts of individuals that were asked to self-isolate that have been reached within 24h. This excludes data pertaining to cases where the individuals that are supposed to self-isolate have not been contacted and may also exclude individuals who have not provided any details of close contacts. Individuals that were asked to self-isolate in response to a positive test in weeks 39 and 40 were affected by the Excel error. The untimely referral caused the contact tracing performance to decline.

**Figure A2:**
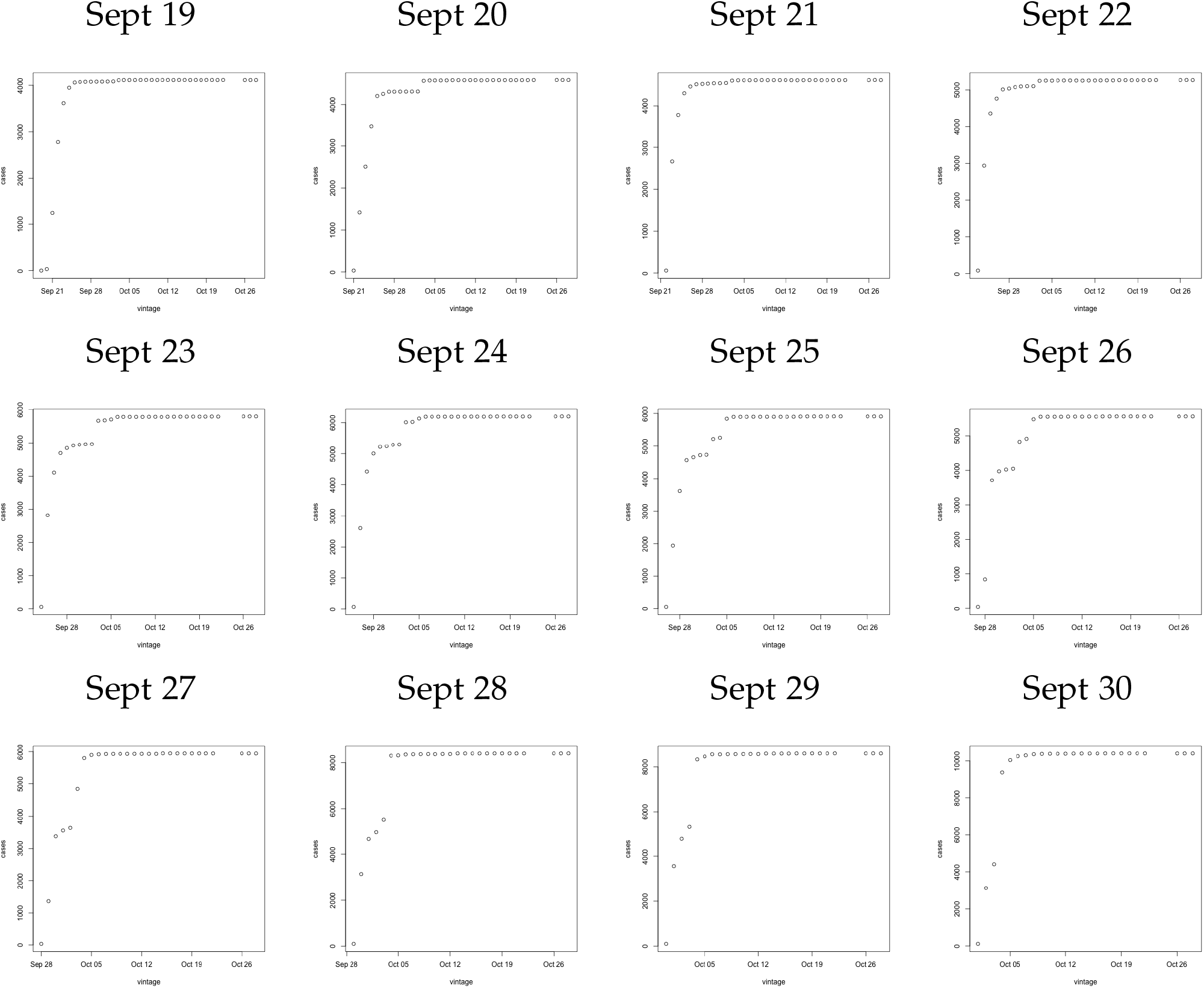
Delayed referral to contact tracing affecting individual cases with positive test result from Sept 20 - Sept 30. **Notes:** Figure plots the number of positive tests on the date indicated in the column head. The vertical axis presents the number of positive cases while the horizontal axis presents the date on which a case count was published. There are notable jumps in the case counts starting Sept 20 due to positive cases not being reported and submitted to contact tracing due to an excel spreadsheet error.

**Figure A3:**
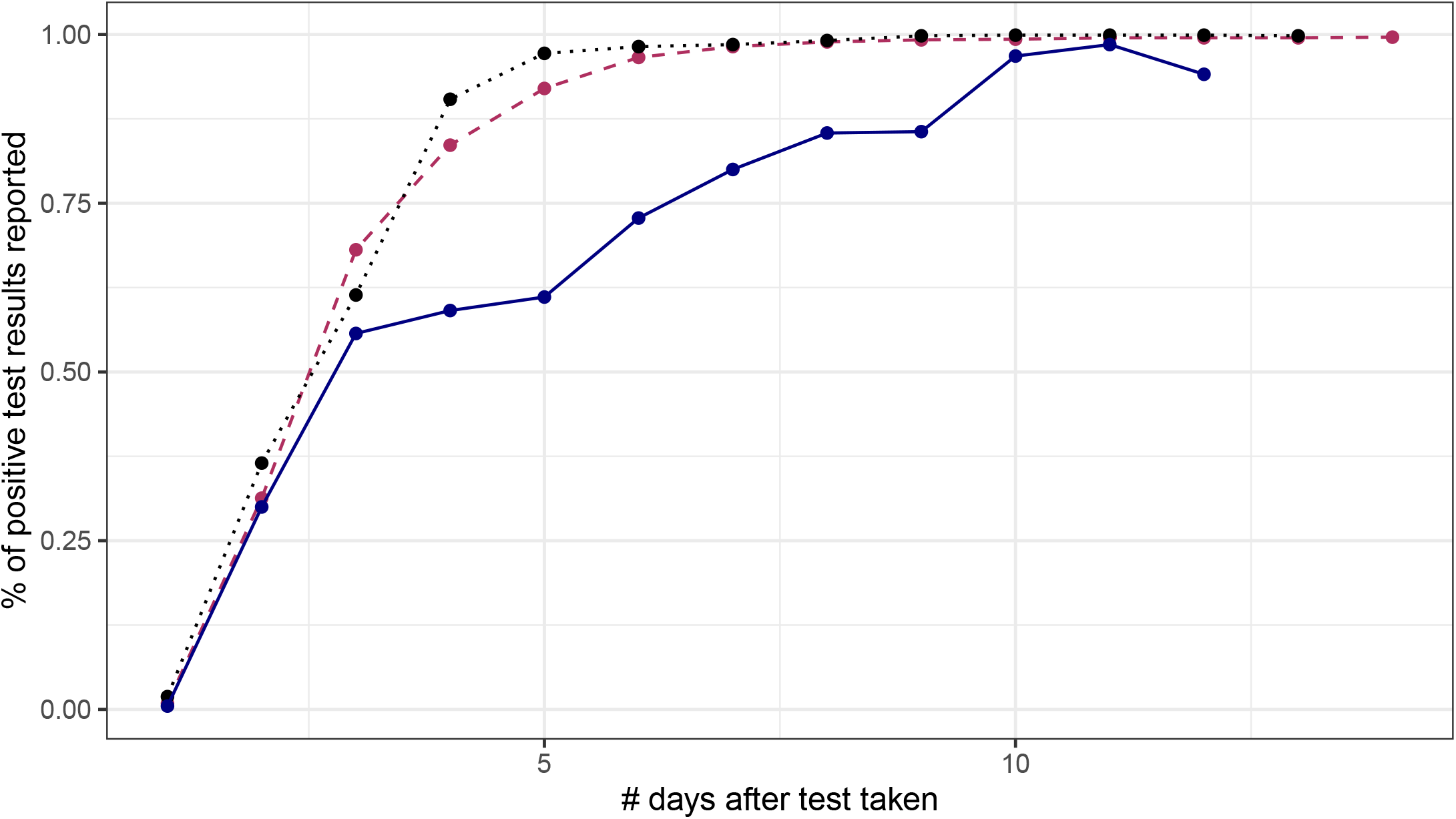
Fraction of positive COVID-19 cases tested on a specific date by reporting delay. **Notes:** Figure plots the share of positive COVID-19 test results that are reported, published and referred to contact tracing as a function of the number of days since the test was taken. The maroon dashed line represents case data from Sept 1 to Sept 20, 2020. On day 5 after the test was taken, on average, 92% of all test results have been published and individuals have been referred to contact tracing. The blue line represents the same curve but for tests performed from Sept 20 to Oct 1st. There are notably fewer positive cases reported and referred to contact tracing as a result of the spreadsheet error. Up to five days after the specimen for a test was taken only 61% of positive test results have been published. The black dotted line presents the same data but for the period from Oct 5 to Oct 15 highlighting this was a temporary glitch.

**Figure A4:**
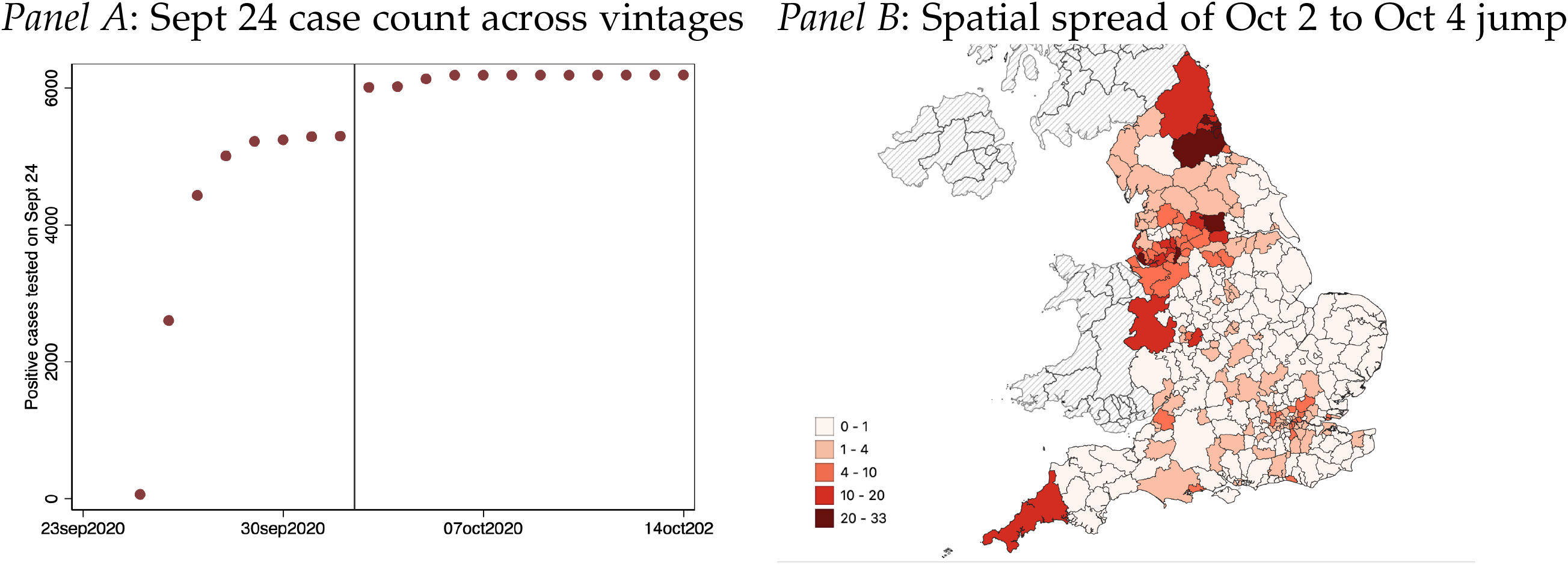
Geographic Signature of Oct 4 upward revision of Sept 24 COVID-19 positive cases across districts. **Notes:** Figure plots the COVID-19 case figures as reported for Sept 24 across different reporting dates in Panel A. of positive COVID-19 test results that are reported, published and referred to contact tracing as a function of the number of days since the test was taken. The maroon dashed line represents case data from Sept 1 to Sept 20, 2020. On day 5 after the test was taken, on average, 92% of all test results have been published and individuals have been referred to contact tracing. The blue line represents the same curve but for tests performed from Sept 20 to Oct 1st. There are notably fewer positive cases reported and referred to contact tracing as a result of the spreadsheet error. Up to five days after the specimen for a test was taken only 61% of positive test results have been published. The black dotted line presents the same data but for the period from Oct 5 to Oct 15 highlighting this was a temporary glitch.

**Figure A5:**
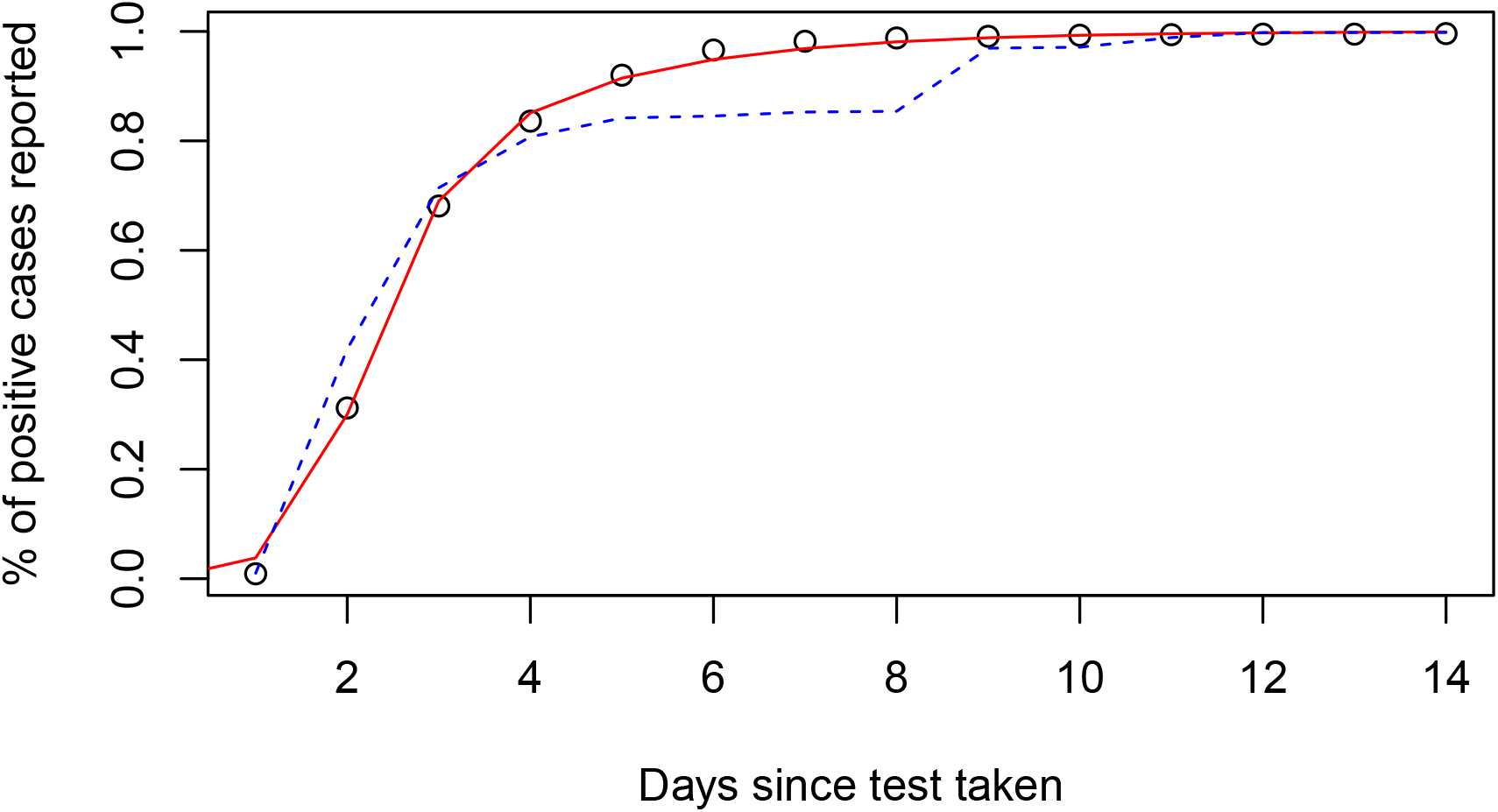
Fitting Evolution of Reported Cases since test date. **Notes:** Figure plots data capturing the share of all positive COVID-19 tests that have been processed, reported and referred to contact tracing as a function of the number of days that have passed since the COVID-19 test was taken on the horizontal axis. The hallow circles refers to the average pattern in the data for Sept 1 to Sept 19. The red line is the one obtained from fitting non-linear least squares of equation 1. The dashed line presents the evolution of the fraction of COVID-19 cases reported and referred to contact tracing for tests taken on Sept 24. The fraction of reported cases jumps nine days after the test was taken which coincides with the upward revision of October 3.

**Figure A6:**
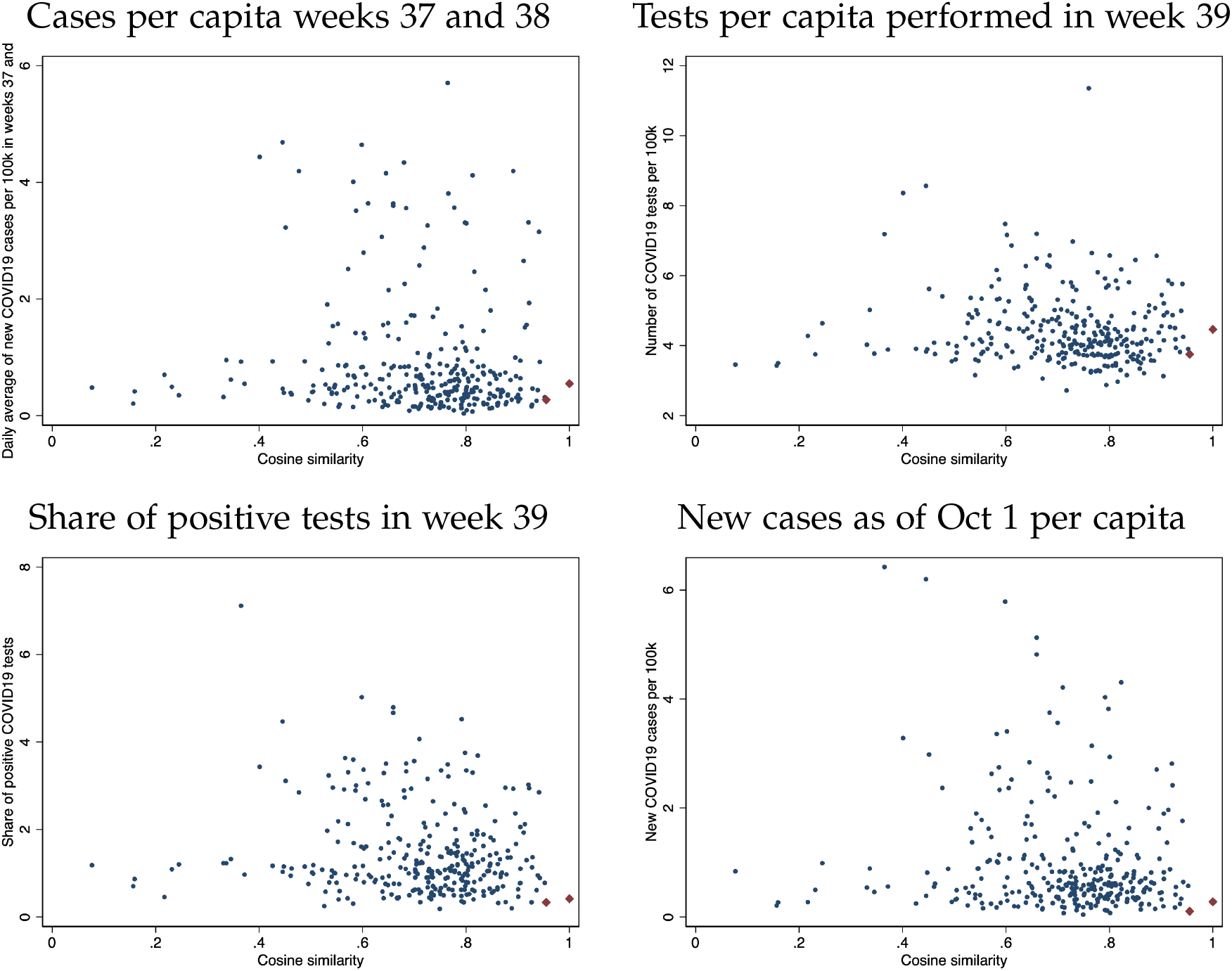
Example Visualisation of Cosine Similarity measure for a pair of districts. **Notes:** Figure plots example of the similarity measure used to construct matched pairs. The cosine similarity measure is plotted along the horizontal axis. The vertical axis presents a subset of features that are included in the cosine similarity measure. The matched pairs are indicated as red diamonds representing two districts that are closest in terms of cosine similarity and form a matched pair. Throughout, the two districts are very similar not just in terms of cosine similarity but also in terms of similarity regarding each individual uni-dimensional measure.

**Figure A7:**
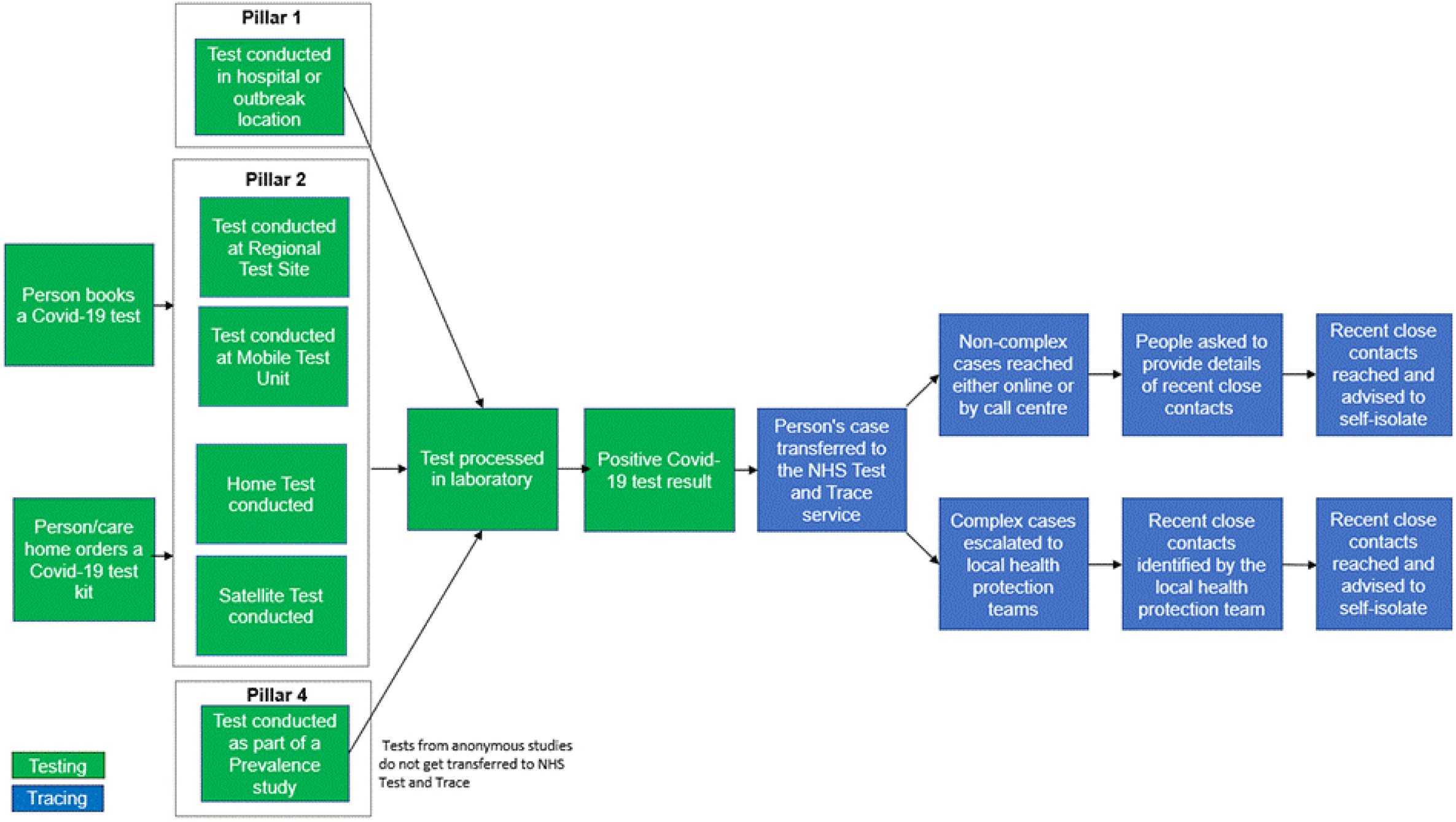
Contact Tracing Flowchart. **Notes:** Figure presents the process activating contact tracing as presented on GOV.UK (2020c).

**Figure A8:**
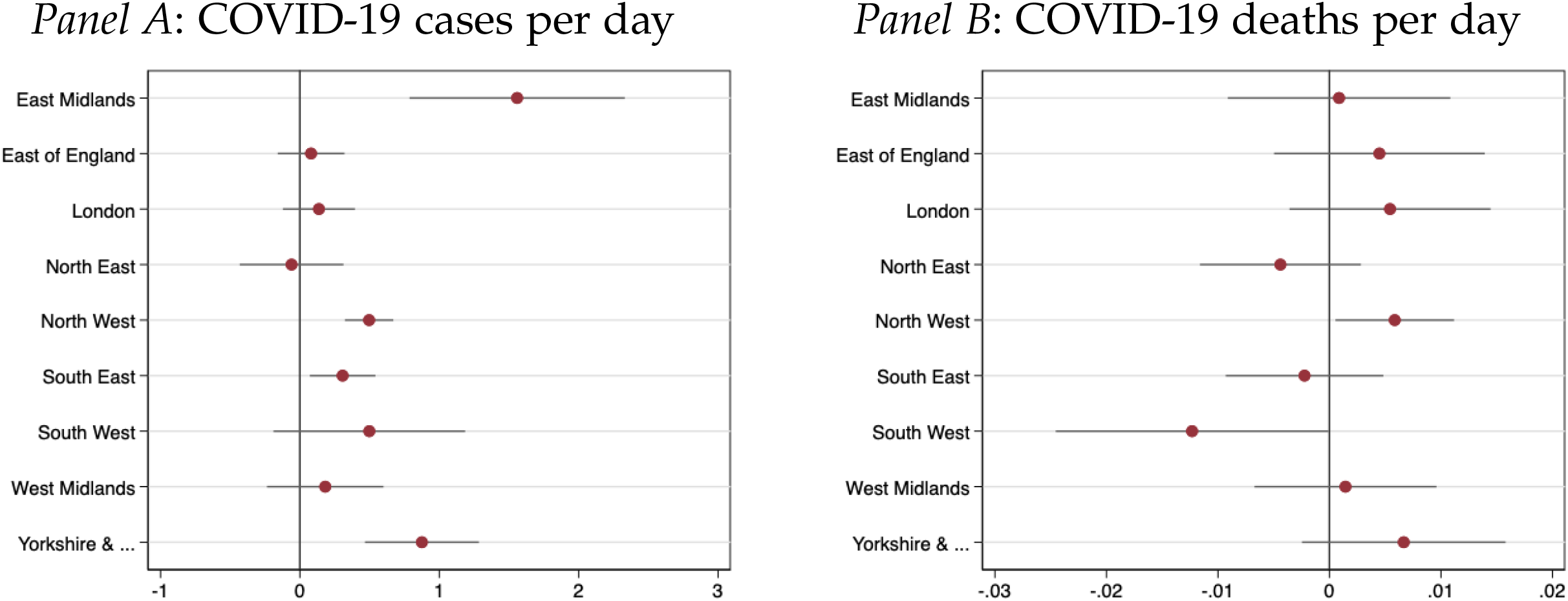
Regional Heterogeneity in Impact of Delayed Contact Tracing Referral on new cases and deaths. **Notes:** Figure plots the impact of delayed referrals to test and trace on subsequent new COVID-19 cases (left panel) and new COVID-19 deaths (right panel). All regressions correspond to the specifications presented in column (1) of Table 2, but allowing the effect to be heterogenous across regions. 90% confidence intervals obtained from clustering standard errors at the district level are indicated.

**Figure A9:**
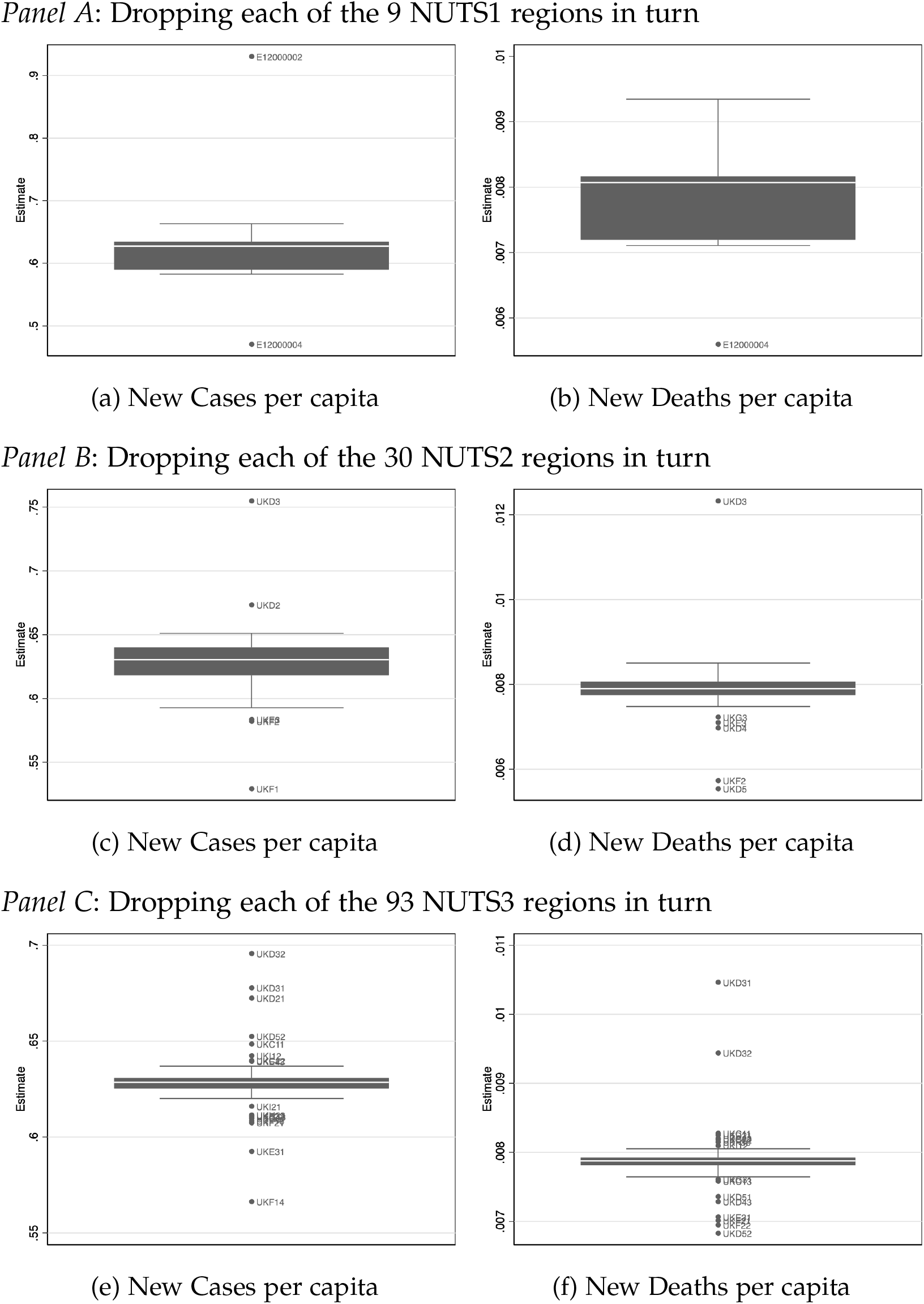
Distribution of point estimates when dropping one region a time. **Notes:** Figures present the distribution of the point estimates obtained when dropping all observations pertaining to one region a time. The estimating regression has as dependent variable either the number of new COVID-19 infections or the number of new COVID-19 deaths after week 40 as recorded in the most recent data version. All regressions control for area fixed effects, time fixed effects and a non-linear time trend in the extent of the local COVID-19 spread measured per capita during weeks 37 and 38. Standard errors are clustered at the district level.

**Figure A10:**
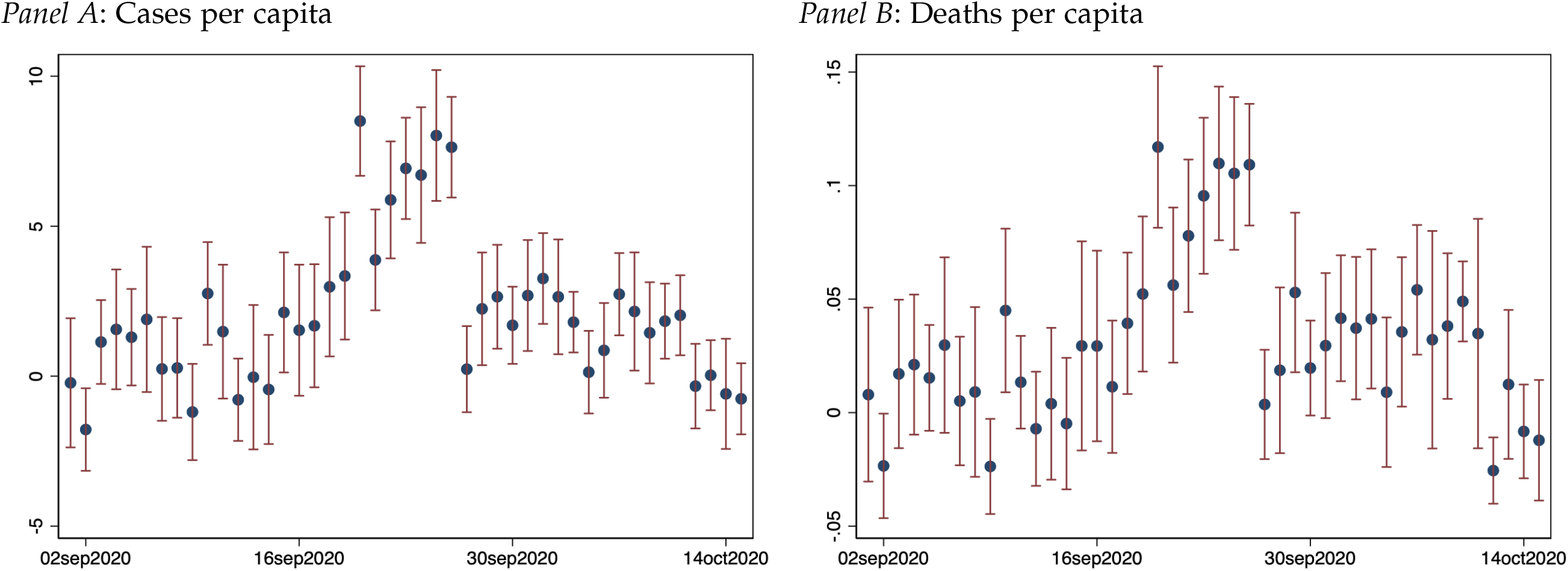
Placebo and actual treatment-effect estimates: non-referred COVID-19 cases and. **Notes:** Figure presents regression estimates of the number of missing cases on subsequent new COVID-19 case growth post calendar week 40 in Panel A and new COVID-19 deaths in Panel B. The number of missing cases on a specific date is computed by measuring, for each date, the number of missing cases is measured as the difference between the case count reported in the most recent data version from November 10, 2020 and the case count published *seven days after* the actual test was taken. That is, the Sept 23 figures represent the gap in reported cases between the Sept 30 version of the case count and the Nov 10, 2020 version of the case count for Sept 23. This implies missing cases affected by the Excel glitch would appear in all data from Sep 20 to Sep 26 as the Excel error was only starting to be rectified from October 3. Data before Sept 20 and after Sept 30 serves as a placebo estimate. Standard errors are clustered at the district level with 90% confidence intervals shown.

**Figure A11:**
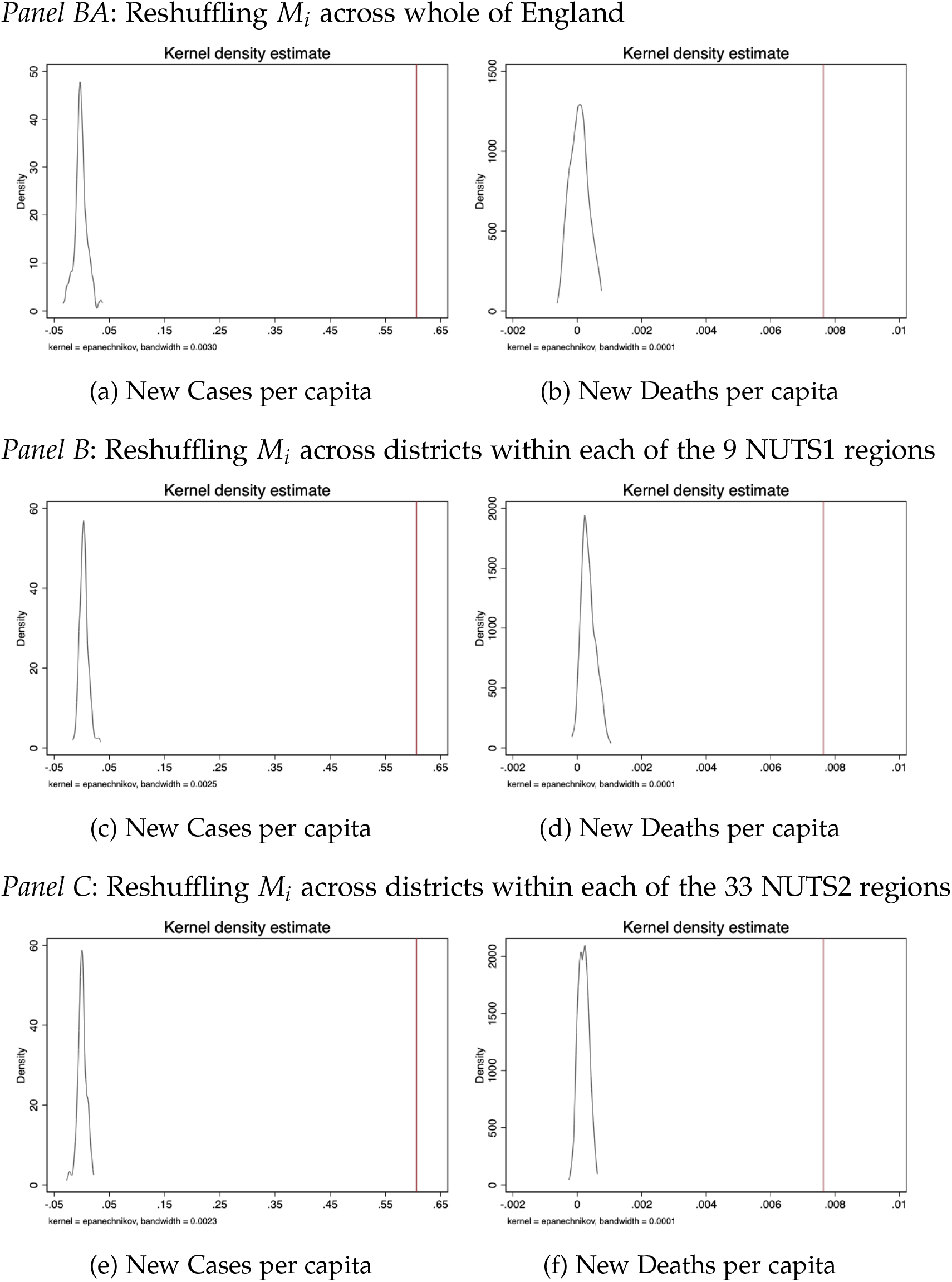
Randomisation inference: reshuffling the district-level exposure measure randomly. **Notes:** Figures present the distribution of point estimates obtained from estimating the main difference-in-difference specification in column (1) of Table 2 when using 100 different reshuffled treatment exposure measure. Reshuffling is either across all districts in England in Panel A; across all districts within each of the 9 NUTS1 regions; across all districts within each of the 33 NUTS2 regions. The kernel density plots the distribution of the point estimates. The vertical line indicates the point estimate obtained when using the actual *M*_*i*_ estimate which corresponds to the point estimates presented in column (1) of Table 2.

**Table A1:**
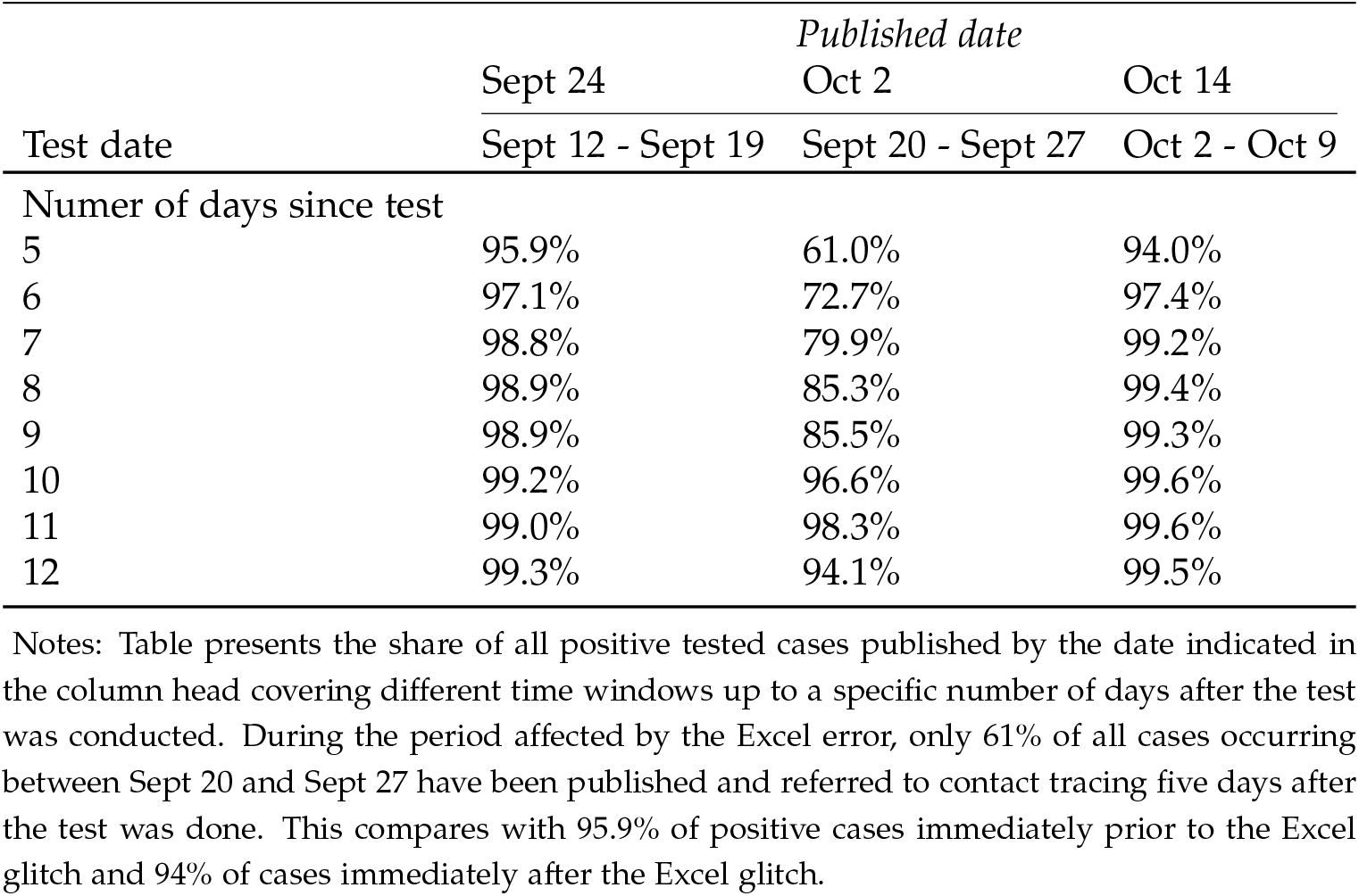
Comparison of usual case count share reported at least five days after a test was taken across different data windows

**Table A2:** Robustness of Impact of non-timely contact tracing on the pandemic progression : Analysis at the Upper Tier Local Authority

|  | *plain difference-in-difference* |  |  | *matched pairs* |  |  |
| --- | --- | --- | --- | --- | --- | --- |
|  | (1) | (2) | (3) | (4) | (5) | (5) |
| *Panel A: New Cases* |  |  |  |  |  |  |
| Post week 39 × Missing cases 20-27 Sept per capita | 0.619***<br>(0.160) | 0.621***<br>(0.157) | 0.635***<br>(0.161) | 0.680***<br>(0.168) | 0.688***<br>(0.176) | 0.776***<br>(0.202) |
| Mean DV | 14.069 | 14.115 | 14.228 | 14.054 | 14.043 | 14.083 |
| Observations | 14378 | 14287 | 14105 | 14014 | 13832 | 13468 |
| Spatial units | 149 | 148 | 146 | 145 | 143 | 139 |
| Additional controls | 1001 | 1911 | 2821 | 7735 | 8554 | 9282 |
| *Panel B: Deaths* |  |  |  |  |  |  |
| Post week 39 × Missing cases 20-27 Sept per capita | 0.010***<br>(0.004) | 0.009**<br>(0.004) | 0.009**<br>(0.004) | 0.011***<br>(0.003) | 0.011***<br>(0.003) | 0.014***<br>(0.003) |
| Mean DV | 0.202 | 0.202 | 0.204 | 0.201 | 0.201 | 0.202 |
| Observations | 14378 | 14287 | 14105 | 14014 | 13832 | 13468 |
| Spatial units | 149 | 148 | 146 | 145 | 143 | 139 |
| Additional controls | 1001 | 1911 | 2821 | 7735 | 8554 | 9282 |
| *Panel C: Tests* |  |  |  |  |  |  |
| Post week 39 × Missing cases 20-27 Sept per capita | 2.934***<br>(0.574) | 2.842***<br>(0.602) | 2.878***<br>(0.652) | 3.599***<br>(0.678) | 3.441***<br>(0.665) | 4.166***<br>(0.894) |
| Mean DV | 311.369 | 311.369 | 311.714 | 311.617 | 311.617 | 311.740 |
| Observations | 14287 | 14287 | 14105 | 13832 | 13832 | 13468 |
| Spatial units | 148 | 148 | 146 | 143 | 143 | 139 |
| Additional controls | 1001 | 1911 | 2821 | 7644 | 8554 | 9282 |
| *Panel D: Positivity rate* |  |  |  |  |  |  |
| Post week 39 × Missing cases 20-27 Sept per capita | 0.001***<br>(0.000) | 0.001***<br>(0.000) | 0.001***<br>(0.000) | 0.001***<br>(0.000) | 0.001***<br>(0.000) | 0.001***<br>(0.000) |
| Mean DV | 0.044 | 0.044 | 0.044 | 0.045 | 0.045 | 0.045 |
| Observations | 13860 | 13860 | 13769 | 13118 | 13118 | 12936 |
| Spatial units | 148 | 148 | 146 | 143 | 143 | 139 |
| Additional controls | 1001 | 1911 | 2821 | 7322 | 8232 | 9051 |
| *Panel E: Contact tracing referral* |  |  |  |  |  |  |
| Post week 39 × Missing cases 20-27 Sept per capita | -0.000<br>(0.001) | -0.000<br>(0.001) | -0.001<br>(0.001) | -0.002*<br>(0.001) | -0.001<br>(0.001) | -0.002*<br>(0.001) |
| Mean DV | 0.633 | 0.633 | 0.633 | 0.633 | 0.633 | 0.633 |
| Observations | 14378 | 14287 | 14105 | 14014 | 13832 | 13468 |
| Spatial units | 149 | 148 | 146 | 145 | 143 | 139 |
| Additional controls | 1001 | 1911 | 2821 | 7735 | 8554 | 9282 |
| **Non-linear time trends in COVID-19 intensity weeks 36-38** |  |  |  |  |  |  |
| Cases per capita deciles x Date | X | X | X | X | X | X |
| Tests per capita deciles x Date |  | X | X |  | X | X |
| Positive test rate deciles x Date |  |  | X |  |  | X |

**Table A3:**
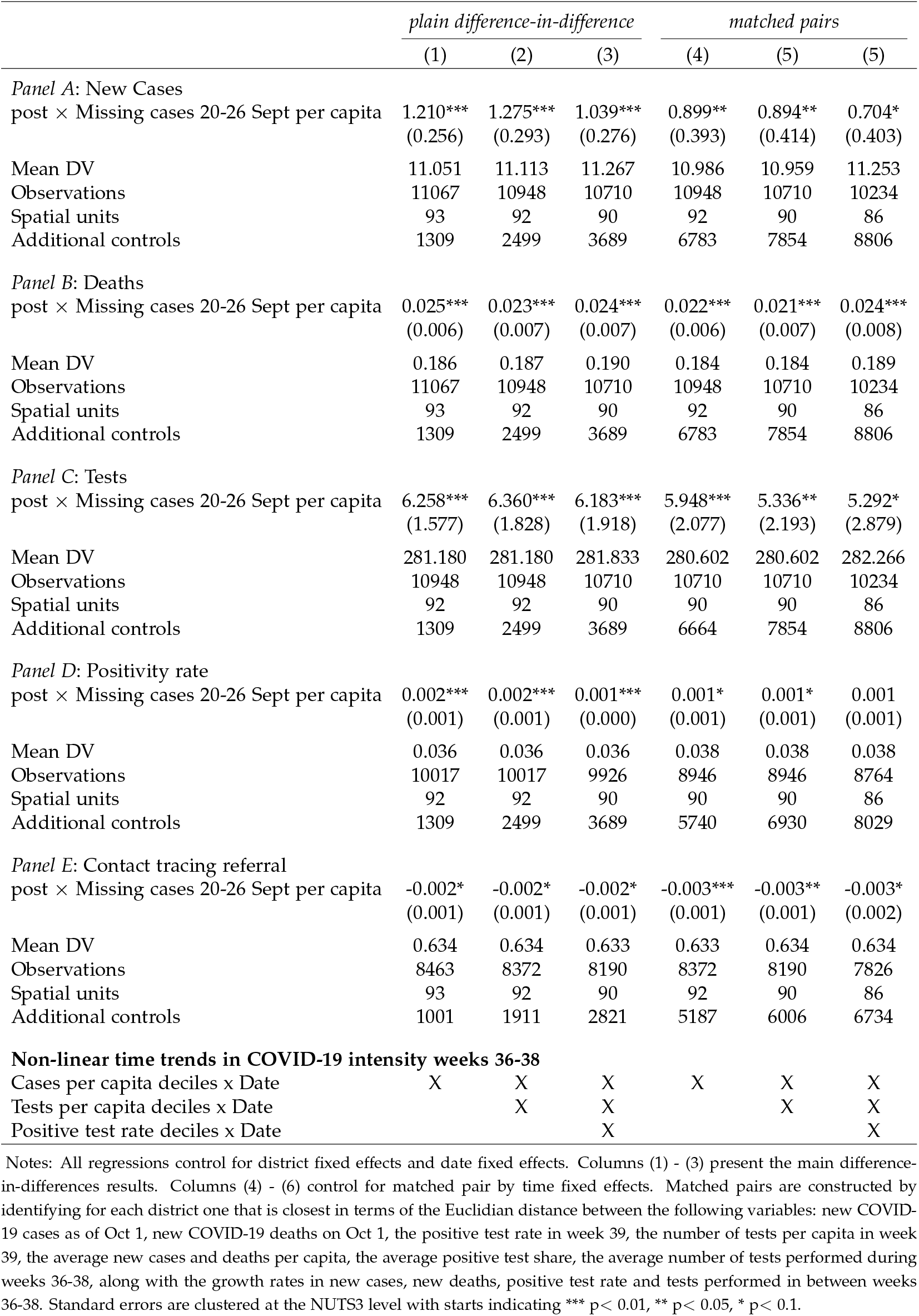
Robustness of Impact of non-timely contact tracing on the pandemic progression : Analysis at the NUTS3 level

|  | *plain difference-in-difference* |  |  | *matched pairs* |  |  |
| --- | --- | --- | --- | --- | --- | --- |
|  | (1) | (2) | (3) | (4) | (5) | (5) |
| *Panel A: New Cases* |  |  |  |  |  |  |
| post × Missing cases 20-26 Sept per capita | 1.210***<br>(0.256) | 1.275***<br>(0.293) | 1.039***<br>(0.276) | 0.899**<br>(0.393) | 0.894**<br>(0.414) | 0.704*<br>(0.403) |
| Mean DV | 11.051 | 11.113 | 11.267 | 10.986 | 10.959 | 11.253 |
| Observations | 11067 | 10948 | 10710 | 10948 | 10710 | 10234 |
| Spatial units | 93 | 92 | 90 | 92 | 90 | 86 |
| Additional controls | 1309 | 2499 | 3689 | 6783 | 7854 | 8806 |
| *Panel B: Deaths* |  |  |  |  |  |  |
| post × Missing cases 20-26 Sept per capita | 0.025***<br>(0.006) | 0.023***<br>(0.007) | 0.024***<br>(0.007) | 0.022***<br>(0.006) | 0.021***<br>(0.007) | 0.024***<br>(0.008) |
| Mean DV | 0.186 | 0.187 | 0.190 | 0.184 | 0.184 | 0.189 |
| Observations | 11067 | 10948 | 10710 | 10948 | 10710 | 10234 |
| Spatial units | 93 | 92 | 90 | 92 | 90 | 86 |
| Additional controls | 1309 | 2499 | 3689 | 6783 | 7854 | 8806 |
| *Panel C: Tests* |  |  |  |  |  |  |
| post × Missing cases 20-26 Sept per capita | 6.258***<br>(1.577) | 6.360***<br>(1.828) | 6.183***<br>(1.918) | 5.948***<br>(2.077) | 5.336**<br>(2.193) | 5.292*<br>(2.879) |
| Mean DV | 281.180 | 281.180 | 281.833 | 280.602 | 280.602 | 282.266 |
| Observations | 10948 | 10948 | 10710 | 10710 | 10710 | 10234 |
| Spatial units | 92 | 92 | 90 | 90 | 90 | 86 |
| Additional controls | 1309 | 2499 | 3689 | 6664 | 7854 | 8806 |
| *Panel D: Positivity rate* |  |  |  |  |  |  |
| post × Missing cases 20-26 Sept per capita | 0.002***<br>(0.001) | 0.002***<br>(0.001) | 0.001***<br>(0.000) | 0.001*<br>(0.001) | 0.001*<br>(0.001) | 0.001<br>(0.001) |
| Mean DV | 0.036 | 0.036 | 0.036 | 0.038 | 0.038 | 0.038 |
| Observations | 10017 | 10017 | 9926 | 8946 | 8946 | 8764 |
| Spatial units | 92 | 92 | 90 | 90 | 90 | 86 |
| Additional controls | 1309 | 2499 | 3689 | 5740 | 6930 | 8029 |
| *Panel E: Contact tracing referral* |  |  |  |  |  |  |
| post × Missing cases 20-26 Sept per capita | -0.002*<br>(0.001) | -0.002*<br>(0.001) | -0.002*<br>(0.001) | -0.003***<br>(0.001) | -0.003**<br>(0.001) | -0.003*<br>(0.002) |
| Mean DV | 0.634 | 0.634 | 0.633 | 0.633 | 0.634 | 0.634 |
| Observations | 8463 | 8372 | 8190 | 8372 | 8190 | 7826 |
| Spatial units | 93 | 92 | 90 | 92 | 90 | 86 |
| Additional controls | 1001 | 1911 | 2821 | 5187 | 6006 | 6734 |
| **Non-linear time trends in COVID-19 intensity weeks 36-38** |  |  |  |  |  |  |
| Cases per capita deciles x Date | X | X | X | X | X | X |
| Tests per capita deciles x Date |  | X | X |  | X | X |
| Positive test rate deciles x Date |  |  | X |  |  | X |

**Table A4:** Robustness of Results to Alternative Functional Forms

|  | *New COVID-19 cases* |  |  | *New COVID-19 deaths* |  |  |
| --- | --- | --- | --- | --- | --- | --- |
|  | (1) | (2) | (3) | (4) | (5) | (5) |
| *Panel A: Main specification* |  |  |  |  |  |  |
| Post week 39 × Missing cases 20-27 Sept per capita | 0.606***<br>(0.111) | 0.619***<br>(0.111) | 0.606***<br>(0.119) | 0.008***<br>(0.003) | 0.008***<br>(0.003) | 0.007**<br>(0.003) |
| Mean DV | 11.790 | 11.848 | 11.906 | 0.183 | 0.184 | 0.185 |
| Observations | 28665 | 28301 | 28119 | 28665 | 28301 | 28119 |
| Spatial units | 315 | 311 | 309 | 315 | 311 | 309 |
| Additional controls | 1001 | 1911 | 2821 | 1001 | 1911 | 2821 |
| *Panel B: Log-Log per capita* |  |  |  |  |  |  |
| Post week 39 × log(Missing cases 20-27 Sept +1 per capita) | 0.239***<br>(0.045) | 0.248***<br>(0.045) | 0.224***<br>(0.048) | 0.045**<br>(0.021) | 0.050**<br>(0.021) | 0.045**<br>(0.023) |
| Mean DV | 1.746 | 1.748 | 1.753 | -0.228 | -0.229 | -0.233 |
| Observations | 28665 | 28301 | 28119 | 28665 | 28301 | 28119 |
| Spatial units | 315 | 311 | 309 | 315 | 311 | 309 |
| Additional controls | 1001 | 1911 | 2821 | 1001 | 1911 | 2821 |
| *Panel C: Levels-Levels* |  |  |  |  |  |  |
| Post week 39 × Missing cases 20-27 Sept | 0.977***<br>(0.139) | 0.967***<br>(0.131) | 0.970***<br>(0.135) | 0.009***<br>(0.003) | 0.009***<br>(0.003) | 0.009***<br>(0.003) |
| Mean DV | 24.169 | 24.361 | 24.505 | 0.346 | 0.348 | 0.350 |
| Observations | 28665 | 28301 | 28119 | 28665 | 28301 | 28119 |
| Spatial units | 315 | 311 | 309 | 315 | 311 | 309 |
| Additional controls | 1001 | 1911 | 2821 | 1001 | 1911 | 2821 |
| *Panel D: Log-Levels per capita* |  |  |  |  |  |  |
| Post week 39 × Missing cases 20-27 Sept per capita | 0.020***<br>(0.004) | 0.022***<br>(0.004) | 0.022***<br>(0.004) | 0.006**<br>(0.003) | 0.007**<br>(0.003) | 0.007**<br>(0.003) |
| Mean DV | 1.746 | 1.748 | 1.753 | -0.228 | -0.229 | -0.233 |
| Observations | 28665 | 28301 | 28119 | 28665 | 28301 | 28119 |
| Spatial units | 315 | 311 | 309 | 315 | 311 | 309 |
| Additional controls | 1001 | 1911 | 2821 | 1001 | 1911 | 2821 |
| *Panel E: Log-Log* |  |  |  |  |  |  |
| Post week 39 × log(Missing cases 20-27 Sept +1) | 0.220***<br>(0.029) | 0.228***<br>(0.029) | 0.222***<br>(0.031) | 0.097***<br>(0.015) | 0.101***<br>(0.016) | 0.104***<br>(0.017) |
| Mean DV | 2.167 | 2.172 | 2.181 | 0.193 | 0.194 | 0.195 |
| Observations | 28665 | 28301 | 28119 | 28665 | 28301 | 28119 |
| Spatial units | 315 | 311 | 309 | 315 | 311 | 309 |
| Additional controls | 1001 | 1911 | 2821 | 1001 | 1911 | 2821 |
| **Non-linear time trends in COVID-19 intensity weeks 36-38** |  |  |  |  |  |  |
| Cases per capita deciles x Date | X | X | X | X | X | X |
| Tests per capita deciles x Date |  | X | X |  | X | X |
| Positive test rate deciles x Date |  |  | X |  |  | X |

Notes: All regressions control for district fixed effects and date fixed effects. Columns (1) - (3) use new daily COVID-19 cases as dependent variable while columns (4) - (6) explore new daily COVID-19 deaths as dependent variable. Standard errors are clustered at the district level with starts indicating \*\*\* p< 0.01, \*\* p< 0.05, \* p< 0.1.

**Table A5:** Robustness of Results to Additional Controls

|  | *New COVID-19 cases* |  |  | *New COVID-19 deaths* |  |  |
| --- | --- | --- | --- | --- | --- | --- |
|  | (1) | (2) | (3) | (4) | (5) | (5) |
| Post week 39 × Missing cases 20-27 Sept per capita | 0.606***<br>(0.119) | 0.443***<br>(0.107) | 0.417***<br>(0.101) | 0.007**<br>(0.003) | 0.006**<br>(0.003) | 0.005**<br>(0.002) |
| Mean DV | 11.906 | 11.906 | 11.991 | 0.185 | 0.185 | 0.185 |
| Observations | 28119 | 28119 | 27755 | 28119 | 28119 | 27755 |
| Spatial units | 309 | 309 | 305 | 309 | 309 | 305 |
| Additional controls | 2730 | 7371 | 7371 | 2730 | 7371 | 7371 |
| Non-linear time trends in COVID-19 intensity weeks 36-38 | X | X | X | X | X | X |
| Non-linear time trends in vector of area controls |  | X | X |  | X | X |
| Google Daily Mobility Scores |  |  | X |  |  | X |

Notes: All regressions control for district fixed effects and date fixed effects. Columns (1) - (3) use new daily COVID-19 cases as dependent variable while columns (4) - (6) explore new daily COVID-19 deaths as dependent variable. Standard errors are clustered at the district level with stars indicating \*\*\* $p < 0.01$ , \*\* $p < 0.05$ , \* $p < 0.1$ .

**Table A6:** Robustness of Results to Alternative Treatment Exposure Measures

|  | *New COVID-19 cases* |  |  | *New COVID-19 deaths* |  |  |
| --- | --- | --- | --- | --- | --- | --- |
|  | (1) | (2) | (3) | (4) | (5) | (6) |
| *Panel A: Main specification* |  |  |  |  |  |  |
| Post week 39 × Missing cases 20-27 Sept per capita | 0.606***<br>(0.111) | 0.619***<br>(0.111) | 0.606***<br>(0.119) | 0.008***<br>(0.003) | 0.008***<br>(0.003) | 0.007**<br>(0.003) |
| Mean DV | 11.790 | 11.848 | 11.906 | 0.183 | 0.184 | 0.185 |
| Observations | 28665 | 28301 | 28119 | 28665 | 28301 | 28119 |
| Spatial units | 315 | 311 | 309 | 315 | 311 | 309 |
| Additional controls | 1001 | 1911 | 2821 | 1001 | 1911 | 2821 |
| *Panel B: Curve fit Sep 20 - Sep 27* |  |  |  |  |  |  |
| Post week 39 × Missing cases Sept/Oct per capita | 0.654***<br>(0.125) | 0.664***<br>(0.126) | 0.641***<br>(0.133) | 0.008***<br>(0.003) | 0.009***<br>(0.003) | 0.008**<br>(0.003) |
| Mean DV | 11.790 | 11.848 | 11.906 | 0.183 | 0.184 | 0.185 |
| Observations | 28665 | 28301 | 28119 | 28665 | 28301 | 28119 |
| Spatial units | 315 | 311 | 309 | 315 | 311 | 309 |
| Additional controls | 1001 | 1911 | 2821 | 1001 | 1911 | 2821 |
| *Panel B: Curve fit Sep 20 - Oct 1* |  |  |  |  |  |  |
| Post week 39 × Missing cases Sept/Oct per capita | 0.442***<br>(0.077) | 0.452***<br>(0.076) | 0.430***<br>(0.076) | 0.005***<br>(0.002) | 0.005***<br>(0.002) | 0.005**<br>(0.002) |
| Mean DV | 11.790 | 11.848 | 11.906 | 0.183 | 0.184 | 0.185 |
| Observations | 28665 | 28301 | 28119 | 28665 | 28301 | 28119 |
| Spatial units | 315 | 311 | 309 | 315 | 311 | 309 |
| Additional controls | 1001 | 1911 | 2821 | 1001 | 1911 | 2821 |
| *Panel D: Fraction Missing cases Sept 20-27* |  |  |  |  |  |  |
| Post week 39 × fracmissed2027sh | 12.705**<br>(5.650) | 12.232*<br>(6.246) | 11.369*<br>(6.656) | 0.037<br>(0.098) | 0.053<br>(0.104) | 0.015<br>(0.111) |
| Mean DV | 11.790 | 11.848 | 11.906 | 0.183 | 0.184 | 0.185 |
| Observations | 28665 | 28301 | 28119 | 28665 | 28301 | 28119 |
| Spatial units | 315 | 311 | 309 | 315 | 311 | 309 |
| Additional controls | 1001 | 1911 | 2821 | 1001 | 1911 | 2821 |
| *Panel E: Missing cases Sept 20-27 high incidence places* |  |  |  |  |  |  |
| Post week 39 × fracmissed2027sh.cens2 | 39.734***<br>(13.521) | 41.164***<br>(14.468) | 45.858***<br>(15.262) | 0.245<br>(0.216) | 0.311<br>(0.223) | 0.324<br>(0.241) |
| Mean DV | 14.130 | 14.212 | 14.212 | 0.210 | 0.211 | 0.211 |
| Observations | 21385 | 21112 | 21112 | 21385 | 21112 | 21112 |
| Spatial units | 235 | 232 | 232 | 235 | 232 | 232 |
| Additional controls | 1001 | 1911 | 2821 | 1001 | 1911 | 2821 |
| **Non-linear time trends in COVID-19 intensity weeks 36-38** |  |  |  |  |  |  |
| Cases per capita deciles × Date | X | X | X | X | X | X |
| Tests per capita deciles × Date |  | X | X |  | X | X |
| Positive test rate deciles × Date |  |  | X |  |  | X |

Notes: All regressions control for district fixed effects and date fixed effects. Columns (1) - (3) use new daily COVID-19 cases as dependent variable while columns (4) - (6) explore new daily COVID-19 deaths as dependent variable. Standard errors are clustered at the district level with stars indicating \*\*\* p < 0.01, \*\* p < 0.05, \* p < 0.1.

**Table A7:** Impact of non-timely contact tracing on the weekly death statistics as reported by the Office of National Statistics by place of death

|  | *plain difference-in-difference* |  |  | *matched pairs* |  |  |
| --- | --- | --- | --- | --- | --- | --- |
|  | (1) | (2) | (3) | (4) | (5) | (5) |
| *Panel A: Weekly COVID-19 Deaths Per Capita* |  |  |  |  |  |  |
| Post week 39 × Missing cases 20-27 Sept per capita | 0.006***<br>(0.002) | 0.007***<br>(0.002) | 0.006***<br>(0.002) | 0.005**<br>(0.002) | 0.006***<br>(0.002) | 0.006***<br>(0.002) |
| Mean DV | 0.079 | 0.080 | 0.081 | 0.079 | 0.080 | 0.081 |
| Observations | 28665 | 28301 | 28119 | 28574 | 28210 | 28028 |
| Spatial units | 315 | 311 | 309 | 314 | 310 | 308 |
| Additional controls | 1001 | 1911 | 2821 | 15288 | 16198 | 17108 |
| *Panel B: Weekly COVID-19 Deaths Recorded in Hospitals Per Capita* |  |  |  |  |  |  |
| Post week 39 × Missing cases 20-27 Sept per capita | 0.006***<br>(0.002) | 0.006***<br>(0.002) | 0.005***<br>(0.002) | 0.006***<br>(0.002) | 0.006***<br>(0.002) | 0.006***<br>(0.002) |
| Mean DV | 0.060 | 0.060 | 0.060 | 0.060 | 0.060 | 0.060 |
| Observations | 28189 | 28189 | 28007 | 28098 | 28098 | 27916 |
| Spatial units | 311 | 311 | 309 | 310 | 310 | 308 |
| Additional controls | 1001 | 1911 | 2821 | 15288 | 16198 | 17101 |
| *Panel C: Weekly COVID-19 Deaths Recorded in Care Homes Per Capita* |  |  |  |  |  |  |
| Post week 39 × Missing cases 20-27 Sept per capita | 0.000<br>(0.001) | 0.001<br>(0.001) | 0.001<br>(0.001) | -0.001<br>(0.001) | -0.001<br>(0.001) | -0.001<br>(0.001) |
| Mean DV | 0.015 | 0.015 | 0.015 | 0.015 | 0.015 | 0.015 |
| Observations | 28224 | 28224 | 28042 | 28133 | 28133 | 27951 |
| Spatial units | 311 | 311 | 309 | 310 | 310 | 308 |
| Additional controls | 1001 | 1911 | 2821 | 15288 | 16198 | 17108 |
| *Panel D: Weekly COVID-19 Deaths Recorded in Other Communal Establishments Per Capita* |  |  |  |  |  |  |
| Post week 39 × Missing cases 20-27 Sept per capita | -0.000<br>(0.000) | -0.000<br>(0.000) | -0.000<br>(0.000) | -0.000<br>(0.000) | -0.000<br>(0.000) | -0.000<br>(0.000) |
| Mean DV | 0.000 | 0.000 | 0.000 | 0.000 | 0.000 | 0.000 |
| Observations | 28259 | 28259 | 28077 | 28168 | 28168 | 27986 |
| Spatial units | 311 | 311 | 309 | 310 | 310 | 308 |
| Additional controls | 1001 | 1911 | 2821 | 15288 | 16198 | 17108 |
| **Non-linear time trends in COVID-19 intensity weeks 36-38** |  |  |  |  |  |  |
| Cases per capita deciles x Date | X | X | X | X | X | X |
| Tests per capita deciles x Date |  | X | X |  | X | X |
| Positive test rate deciles x Date |  |  | X |  |  | X |

## Footnotes

1 See, e.g., Guardian (2020); Telegraph (2020); Mirror (2020).

2 Available at https://coronavirus.data.gov.uk/.

3 The authors have launched a public FOIA request to request more granular data. The FOIA request can be accessed here: https://www.whatdotheyknow.com/request/nhs_test_and_trace_statistics_re.

4 The FOIA is in the public domain on https://www.whatdotheyknow.com/request/regiona_l_breakdown_of_cases_not.

## References

Afzal, Ibrahim, Raheema Abdul Raheem, Nazla Rafeeq, and Sheena Moosa, “Contact tracing for containment of novel coronavirus disease (COVID-19) in the early phase of the epidemic in the Maldives,” Asia Pacific Journal of Public Health, 2020, p. 1010539520956447.

Altmann, Samuel, Luke Milsom, Hannah Zillessen, Raffaele Blasone, Frederic Gerdon, Ruben Bach, Frauke Kreuter, Daniele Nosenzo, Séverine Toussaert, and Johannes Abeler, “Acceptability of app-based contact tracing for COVID-19: Cross-country survey study,” JMIR mHealth and uHealth, 2020, 8 (8), e19857.

Anderson, Roy M, Hans Heesterbeek, Don Klinkenberg, and T sDéirdre Hollingsworth, “How will country-based mitigation measures influence the course of the COVID-19 epidemic?,” The Lancet, 2020, 395 (10228), 931–934.

BBC, “Covid: Test error ‘should never have happened’ - Hancock,” 2020, https://www.bbc.com/news/uk-54422505 (Accessed: Nov 13, 2020).

Cho, Hyunghoon, Daphne Ippolito, and Yun William Yu, “Contact tracing mobile apps for COVID-19: Privacy considerations and related trade-offs,” arXiv preprint arXiv:2003.11511,2020.

Clark, Eva, Elizabeth Y Chiao, and E Susan Amirian, “Why contact tracing efforts have failed to curb coronavirus disease 2019 (covid-19) transmission in much of the united states,” Clinical Infectious Diseases, 2020.

Conway, Ed, “Coronavirus: More than 1,000 consultants from Deloitte on Test and Trace programme,” Sky News, 2020, https://news.sky.com/story/coronavirus-more-than-1-000-consultants-from-deloitte-on-test-and-trace-programme-12099127 (Accessed: Nov 13, 2020).

Cowling, Benjamin J and Wey Wen Lim, “They’ve contained the coronavirus. Here’s how,” New York Times, 2020, 13.

Desvars-Larrive, Amelie, Elma Dervic, Nils Haug, Thomas Niederkrotenthaler, Jiaying Chen, Anna Di Natale, Jana Lasser, Diana S Gliga, Alexandra Roux, Abhijit Chakraborty et al., “A structured open dataset of government interventions in response to COVID-19,” medRxiv, 2020.

Fenner, Frank, Donald Ainslie Henderson, Isao Arita, Zdenek Jezek, Ivan D Ladnyi et al., Smallpox and its eradication, Vol. 6, World Health Organization Geneva, 1988.

Ferguson, Neil, Daniel Laydon, Gemma Nedjati-Gilani, Natsuko Imai, Kylie Ainslie, Marc Baguelin, Sangeeta Bhatia, Adhiratha Boonyasiri, Zulma Cucunubá, Gina Cuomo-Dannenburg et al., “Report 9: Impact of nonpharmaceutical interventions (NPIs) to reduce COVID19 mortality and health-care demand,” Imperial College London, 2020, 10, 77482.

Fetzer, Thiemo, “Subsidizing the spread of COVID19: Evidence from the UK’s Eat-Out-to-Help-Out scheme,” CAGE Working Paper, 2020, (517).

Fetzer, Thiemo, Marc Witte, Lukas Hensel, Jon Jachimowicz, Johannes Haushofer, Andriy Ivchenko, Stefano Caria, Elena Reutskaja, Christopher Roth, Stefano Fiorin et al., “Global Behaviors and Perceptions in the COVID-19 Pandemic,” 2020.

Google, “Google COVID-19 Community Mobility Reports.,” 2020, https://www.google.com/covid19/mobility/ (Accessed: Nov 13, 2020).

GOV.UK, “COVID-19 testing data: methodology note,” 2020, https://www.gov.uk/government/publications/coronavirus-covid-19-testing-data-methodology/covid-19-testing-data-methodology-note (Accessed: Nov 13, 2020).

GOV.UK, “NHS Test and Trace (England) and coronavirus testing (UK) statistics,” 2020, https://www.gov.uk/government/publications/nhs-test-and-trace-england-and-coronavirus-testing-uk-statistics-22-october-to-28-october.

GOV.UK, “NHS Test and Trace statistics (England): methodology,” 2020, https://www.gov.uk/government/publications/nhs-test-and-trace-statistics-england-methodology/nhs-test-and-trace-statistics-england-methodology (Accessed: Nov 13, 2020).

GOV.UK, “PHE statement on delayed reporting of COVID-19 cases,” 2020, https://www.gov.uk/government/news/phe-statement-on-delayed-reporting-of-covid-19-cases (Accessed: Nov 13, 2020).

GOV.UK, “Summary of the effectiveness and harms of different non-pharmaceutical interventions, 21 September 2020,” 2020, https://www.gov.uk/government/publications/summary-of-the-effectiveness-and-harms-of-different-non-pharmaceutical-interventions-16-september-2020 (Accessed: Nov 13, 2020).

Grantz, Kyra H, Elizabeth C Lee Lucy D’Agostino McGowan, Kyu Han Lee C Jessica E Metcalf, Emily S Gurley, and Justin Lessler, “Maximizing and evaluating the impact of test-trace-isolate programs,” medRxiv, 2020.

Guardian, The, “Covid: how Excel may have caused loss of 16,000 test results in England,” 2020, https://www.theguardian.com/politics/2020/oct/05/how-excel-may-have-caused-loss-of-16000-covid-tests-in-england (Accessed: Nov 13, 2020).

Hakak, Saqib, Wazir Zada Khan, Muhammad Imran, Kim-Kwang Raymond Choo, and Muhammad Shoaib, “Have you been a victim of COVID-19-related cyber incidents? Survey, taxonomy, and mitigation strategies,” IEEE Access, 2020, 8, 124134–124144.

Kendall, Michelle, Luke Milsom, Lucie Abeler-Dörner, Chris Wymant, Luca Ferretti, Mark Briers, Chris Holmes, David Bonsall, Johannes Abeler, and Christophe Fraser, “Epidemiological changes on the Isle of Wight after the launch of the NHS Test and Trace programme: a preliminary analysis,” The Lancet Digital Health, 2020.

Klinkenberg, Don, Christophe Fraser, and Hans Heesterbeek, “The effectiveness of contact tracing in emerging epidemics,” PloS one, 2006, 1 (1), e12.

Kretzschmar, Mirjam E, Ganna Rozhnova, Martin Bootsma, Michiel E van Boven, Janneke van de Wijgert, and Marc Bonten, “Time is of the essence: impact of delays on effectiveness of contact tracing for COVID-19,” medRxiv, 2020.

Martin CJ Bootsma, Michiel van Boven, Janneke HHM van de Wijgert, and Marc JM Bonten, “Impact of delays on effectiveness of contact tracing strategies for COVID-19: a modelling study,” The Lancet Public Health, 2020, 5 (8), e452–e459.

Kucharski, Adam J, Petra Klepac, Andrew Conlan, Stephen M Kissler, Maria Tang, Hannah Fry, Julia Gog, John Edmunds, CMMID COVID-19 Working Group et al., “Effectiveness of isolation, testing, contact tracing and physical distancing on reducing transmission of SARS-CoV-2 in different settings,” medRxiv, 2020.

Li, Jinfeng and Xinyi Guo, “COVID-19 Contact-tracing Apps: A Survey on the Global Deployment and Challenges,” arXiv preprint arXiv:2005.03599, 2020.

Lu, Ning, Kai-Wen Cheng, Nafees Qamar, Kuo-Cherh Huang, and James A Johnson, “Weathering COVID-19 storm: successful control measures of five Asian countries,” American journal of infection control, 2020, 48 (7), 851–852.

Manthorpe, Roland, “Coronavirus: Untrained staff take on contact-tracing jobs as Test and Trace struggles to cope with rise in cases,” Sky News, 2020, https://news.sky.com/story/coronavirus-untrained-staff-take-on-contact-tracing-jobs-as-test-and-trace-struggles-to-cope-with-rise-in-cases-12110519 (Accessed: Nov 13, 2020).

Mirror, “16,000 coronavirus tests went missing because of Excel spreadsheet blunder,” 2020, https://www.mirror.co.uk/news/politics/16000-coronavirus-tests-went-missing-22794820 (Accessed: Nov 13, 2020).

Mueller, Benjamin and Jane Bradley, “England’s ‘World Beating’ System to Track the Virus Is Anything But,” The New York Times, 2020, https://www.nytimes.com/2020/06/17/world/europe/uk-contact-tracing-coronavirus.html (Accessed: Nov 13, 2020).

NHS, “If you’re told to self-isolate by NHS Test and Trace or the NHS COVID-19 app,” 2020, https://www.nhs.uk/conditions/coronavirus-covid-19/testing-and-tracing/nhs-test-and-trace-if-youve-been-in-contact-with-a-person-who-has-coronavirus/ (Accessed: Nov 13, 2020).

ONS, “Death registrations and occurrences by local authority and health board,” 2020, https://www.ons.gov.uk/peoplepopulationandcommunity/healthandsocialcare/causesofdeath/datasets/ (Accessed: Nov 13, 2020).

Park, YJ, YJ Choe, O Park, SY Park, YM Kim, J Kim et al., “COVID-19 National Emergency Response Center, Epidemiology and Case Management Team. Contact tracing during coronavirus disease outbreak, South Korea, 2020,” Emerg Infect Dis, 2020, 26 (10).

Rubin, G James, Louise E Smith, GJ Melendez-Torres, and Lucy Yardley, “Im-proving adherence to ‘test, trace and isolate’,” Journal of the Royal Society of Medicine, 2020, 113 (9), 335–338.

Simko, Lucy, Ryan Calo, Franziska Roesner, and Tadayoshi Kohno, “COVID-19 Contact Tracing and Privacy: Studying Opinion and Preferences,” arXiv preprint arXiv:2005.06056, 2020.

Steer, Nicola C., Paul M. Ramsay, and Miguel Franco, “nlstimedist: An R package for the biologically meaningful quantification of unimodal phenology distributions,” Methods in Ecology and Evolution, nov 2019, 10 (11), 1934–1940.

Steinhauer, Jennifer and Abby Goodnough, “Contact Tracing is Failing in Many States. Here’s Why.,” The New York Times, 2020, https://www.nytimes.com/2020/07/31/health/covid-contact-tracing-tests.html (Accessed: Nov 13, 2020).

Telegraph, The, “The ‘Excel error’ that led to 16,000 missing coronavirus cases,” 2020, https://www.telegraph.co.uk/technology/2020/10/05/excel-errorled-16000-missing-coronavirus-cases/ (Accessed: Nov 13, 2020).

Webster, Rebecca K, Samantha K Brooks, Louise E Smith, Lisa Woodland, Simon Wessely, and G James Rubin, “How to improve adherence with quarantine: Rapid review of the evidence,” Public Health, 2020.

WHO, “Contact tracing in the context of COVID-19,” 2020, https://www.who.int/publications/i/item/contact-tracing-in-the-context-of-covid-19 (Accessed: Nov 13, 2020).

Yilmazkuday, Hakan, “Stay-at-home works to fight against COVID-19: international evidence from Google mobility data,” Available at SSRN 3571708, 2020.

